# Transcranial direct current stimulation (tDCS) combined with cognitive training in adolescent boys with ADHD: a double-blind, randomised, sham-controlled trial

**DOI:** 10.1101/2020.12.07.20245456

**Authors:** Samuel J. Westwood, Marion Criaud, Sheut-Ling Lam, Steve Lukito, Sophie Wallace-Hanlon, Olivia S. Kowalczyk, Afroditi Kostara, Joseph Mathew, Deborah Agbedjro, Bruce E. Wexler, Roi Cohen Kadosh, Philip Asherson, Katya Rubia

**Affiliations:** Department of Child & Adolescent Psychiatry, King’s College London, UK; School of Psychology, University of Surrey, UK; Department of Neuroimaging, King’s College London, UK; Department of Biostatistics, King’s College London, UK; Department of Psychiatry, Yale University School of Medicine, USA; Department of Experimental Psychology, University of Oxford, UK; Social Genetic & Developmental Psychiatry, King’s College London, UK

**Author notes:** Corresponding author: Dr Samuel J. Westwood, Department of Child and Adolescent Psychiatry PO46, Social, Genetic and Developmental Psychiatry (SGDP) Centre, Institute of Psychiatry, Psychology and Neuroscience, King’s College London, 16 De Crespigny Park, London, SE5 8AF, UK.

**Keywords:** tDCS, ADHD, treatment, randomised controlled trial

## Abstract

**Background:** Transcranial direct current stimulation (tDCS) could be a side-effect free alternative to psychostimulants in Attention-Deficit/Hyperactivity Disorder (ADHD). Although there is limited evidence for clinical and cognitive effects, most studies were small, single-session, and stimulated left dorsolateral prefrontal cortex (dlPFC). No sham-controlled study has stimulated right inferior frontal cortex (rIFC), which is the most consistently under-functioning region in ADHD, with multiple sessions of anodal tDCS combined with cognitive training (CT) to enhance effects.

**Objective/Hypothesis:** To investigate clinical and cognitive effects of multi-session anodal tDCS over rIFC combined with CT in a double-blind, randomised, sham-controlled trial (RCT).

**Methods:** 50 boys with ADHD (10-18 years) received 15 weekday sessions of anodal or sham tDCS over rIFC combined with CT (20mins, 1mA). ANCOVA, adjusting for baseline measures, age, and medication status, tested group differences in clinical and ADHD-relevant executive functions at posttreatment and after 6-months.

**Results:** ADHD-Rating Scale, Conners ADHD Index, and adverse effects were significantly lower at post-treatment after sham relative to real tDCS. No other effects were significant.

**Conclusions:** This rigorous multi-session RCT of tDCS over the rIFC in ADHD combined with CT, showed no evidence of improvement of ADHD symptoms or cognitive performance. Findings extend limited meta-analytic evidence of cognitive and clinical effects in ADHD after 1-5 tDCS sessions over mainly left dlPFC. Given that tDCS is commercially and clinically available, the findings are important as they suggest that rIFC stimulation may not be indicated as a neurotherapy for cognitive or clinical remediation for ADHD

**Highlights:**

- tDCS has been suggested as an alternative treatment for ADHD
- We combined 15-session anodal tDCS over the rIFC with cognitive training in ADHD children
- Real versus sham tDCS showed no cognitive or symptom improvements
- Conversely, real tDCS showed lower ADHD symptoms and higher adverse effects
- Multi-session tDCS of rIFC shows no clinical or cognitive benefits in ADHD

## INTRODUCTION

Attention-Deficit/Hyperactivity Disorder (ADHD) is characterised by persisting, age-inappropriate, and impairing symptoms of inattention and/or impulsivity-hyperactivity [1]. ADHD is also associated with deficits in executive functions (EF), most prominently in motor and interference inhibition, sustained attention and vigilance, switching, working memory, and timing [2,3]. Neuroimaging studies indicate delayed structural and functional brain maturation [4,5], and consistent underactivation in predominantly right inferior frontal (rIFC), dorsolateral prefrontal (dlPFC), and anterior cingulate cortices, as well as striatal, parietal and cerebellar regions during cognitive control, timing, and attention tasks [3,6–9].

Psychostimulants are the gold-standard treatment for improving ADHD symptoms, but have associated side-effects [10], poor adherence in adolescence [11,12], and are not indicated for all individuals with ADHD [10]. Evidence of longer-term efficacy is also limited [12,13], possibly due to brain adaptation [14]. Meta-analyses indicate small to moderate efficacy of behavioural therapies, cognitive training (CT), neurofeedback, or dietary interventions on ADHD symptoms [15]. Non-invasive brain stimulation techniques, however, are promising given their potential to stimulate key dysfunctional brain regions associated with ADHD, with potentially longer-term neuroplastic effects that drugs cannot offer [3,16–19]. Transcranial direct current stimulation (tDCS) is particularly well-suited for paediatric populations as it is user-friendly, well tolerated with minimal side effects [20], and is cheaper relative to other techniques, such as transcranial magnetic stimulation [21].

In tDCS, a weak direct electrical current is delivered through two electrodes placed on the scalp (one anode, one cathode), generating subthreshold, polarity-dependent shifts in resting membrane potentials in underlying brain regions. The resulting net increase (predominantly under the anode) or decrease (predominantly under the cathode) in neuronal excitability leads to modulation of the neuronal network [22], with neurophysiological effects persisting after stimulation, presumably by potentiating mediators of practice-dependent synaptic plasticity, including GABA, glutamate [23,24], dopamine, and noradrenaline [25– 27].

Evidence of cognitive performance and clinical improvement following tDCS is, however, limited. Two meta-analyses of tDCS studies stimulating mainly the left dlPFC in 1-5 sessions in children and adolescents with ADHD indicate a modest improvement in inhibitory control measures [28,29] with the later one showing non-significant improvement on processing speed and inhibitory measures with no effects on attention measures [19]. Only two sham-controlled studies tested ADHD symptoms, reporting improvement in inattentive symptoms, but not impulsivity/hyperactivity, immediately, one [30,31], and two weeks [31] after anodal tDCS over left or right dlPFC.

There is evidence that tDCS effects can be enhanced when combined with CT by functionally priming the brain regions that mediate the cognitive function being trained [32– 35]. Multi-session anodal tDCS combined with CT has also been shown to elicit longer-term cognitive improvements in healthy controls for up to one [36,37], nine [38], or 12 months [34], and clinical improvements in psychiatric disorders for at least one month [39–41], suggesting longer-term neuroplastic effects.

Most studies in ADHD have targeted the left dlPFC. However, one of the most consistent findings of meta-analyses of functional magnetic resonance imaging (fMRI) studies is the underactivation of rIFC during tasks of cognitive control [3,7–9,42]. The rIFC is a cognitive control “hub” region, playing a key role in cognitive control [43,44], sustained attention [45–47], and timing networks [48], mediating key functions of impairment in ADHD [2,49]. The rIFC dysfunction has also been shown to be disorder-specific to ADHD compared to several neurodevelopmental disorders in comparative fMRI meta-analyses of cognitive control [3,8,9]. Further, its upregulation is the most consistent effect of single-dose and longer-term psychostimulant medication [9,50].

Only three published studies applied tDCS over rIFC in ADHD, in 1 or 5 sessions. One found improvements after anodal relative to sham tDCS in Flanker task errors and intrasubject response variability in 21 adolescents with ADHD [51], but no effect in 14 children with ADHD on a combined n-back and Stop task with high definition (HD) or conventional anodal tDCS [52]. In 20 undiagnosed high-school students with ADHD symptoms, anodal compared to sham tDCS improved Go accuracy in a Go/No-Go (GNG) task but no other inhibitory or Stroop task measures [53]. To our knowledge, no sham-controlled study so far has measured clinical and cognitive effects of multi-session anodal tDCS over rIFC in combination with CT in patients with ADHD

In this double-blind, sham-controlled, randomised controlled trial (RCT), 50 children and adolescents with ADHD were administered 15 consecutive weekday sessions of anodal or sham tDCS over rIFC combined with CT of EF typically impaired in ADHD. The primary outcome measures were parent-rated clinical symptoms and cognitive performance in an inhibition and a sustained-attention task. Secondary outcome measures were other clinical, safety, and EF measures.

Given some evidence of clinical and cognitive improvements with anodal tDCS over the dlPFC [30,31] or rIFC [51,53] and prolonged effects when tDCS is paired with CT, we hypothesised that, compared to sham, at posttreatment, multi-session anodal tDCS of rIFC with multi-EF training would show greater improvement in ADHD symptoms and/or performance on EF tasks mediated by rIFC and targeted by the CT. We also hypothesised that clinical and EF task improvement would persist 6-months after posttreatment. Finally, we hypothesised no side or adverse effects.

## MATERIALS AND METHODS

### Trial Design

This pre-registered (ISRCTN: 48265228), double-blind, sham-controlled, parallel RCT compared multi-session anodal versus sham tDCS over the rIFC combined with multi-EF training. Randomisation to stimulation group was stratified by age (10 to 14.5 and 14.5 to 18 years) and medication status (naïve and non-naïve) using Smith randomisation [54,55] conducted by Innosphere Ltd (Haifa, Israel). Experimenters, participants, and parents/caregivers were blind to stimulation group. This trial received local research ethics committee approval (REC ID: 17/LO/0983) and was conducted in accordance with the Declaration of Helsinki and Consolidated Standards of Reporting Trials (CONSORT) guidelines [56].

### Participants

Fifty male participants (10 to 18 years) were recruited from South London clinics, social media, and parent support groups from February 2018 to 2020. All participants had a clinical DSM-5 diagnosis of ADHD established by a clinician, and validated by the Schedule of Affective Disorders and Schizophrenia for School-Age Children-Present and Lifetime version (K-SADS-PL) [57] and the Conners 3rd Edition–Parent (Conners 3-P, cut-off t-score > 60) [58]. Autism spectrum disorders (ASD) was excluded using a combination (both required) of both the parent-rated Social Communication Questionnaire (SCQ, cut-off > 17) [59] and the pro-social scale of the Strengths & Difficulties Questionnaire (SDQ, cut-off < 5) [60]; for participants falling outside these criteria, the absence of ASD was confirmed by their clinician. Further exclusion criteria were IQ□below 80 (Wechsler abbreviated scale of intelligence, WASI-II) [61], history of alcohol or substance abuse, neurological illness, comorbid major psychiatric disorders as established by the K-SADS-PL or clinical diagnoses (except for Conduct Disorder [CD]/Oppositional Defiant Disorder [ODD]), contraindications to tDCS (e.g., metallic implants [except dental braces], previous neurosurgical procedures, history of migraine, diseased/damaged skin on the scalp), treated with drugs that lower seizure thresholds (e.g., antipsychotics, tricyclic antidepressants), and an inability to provide consent from the legal caregiver for children under 16-years or from participants over 16-years (Figure 1). Participants received £540 plus travel expenses for participating.

**Figure 1.**
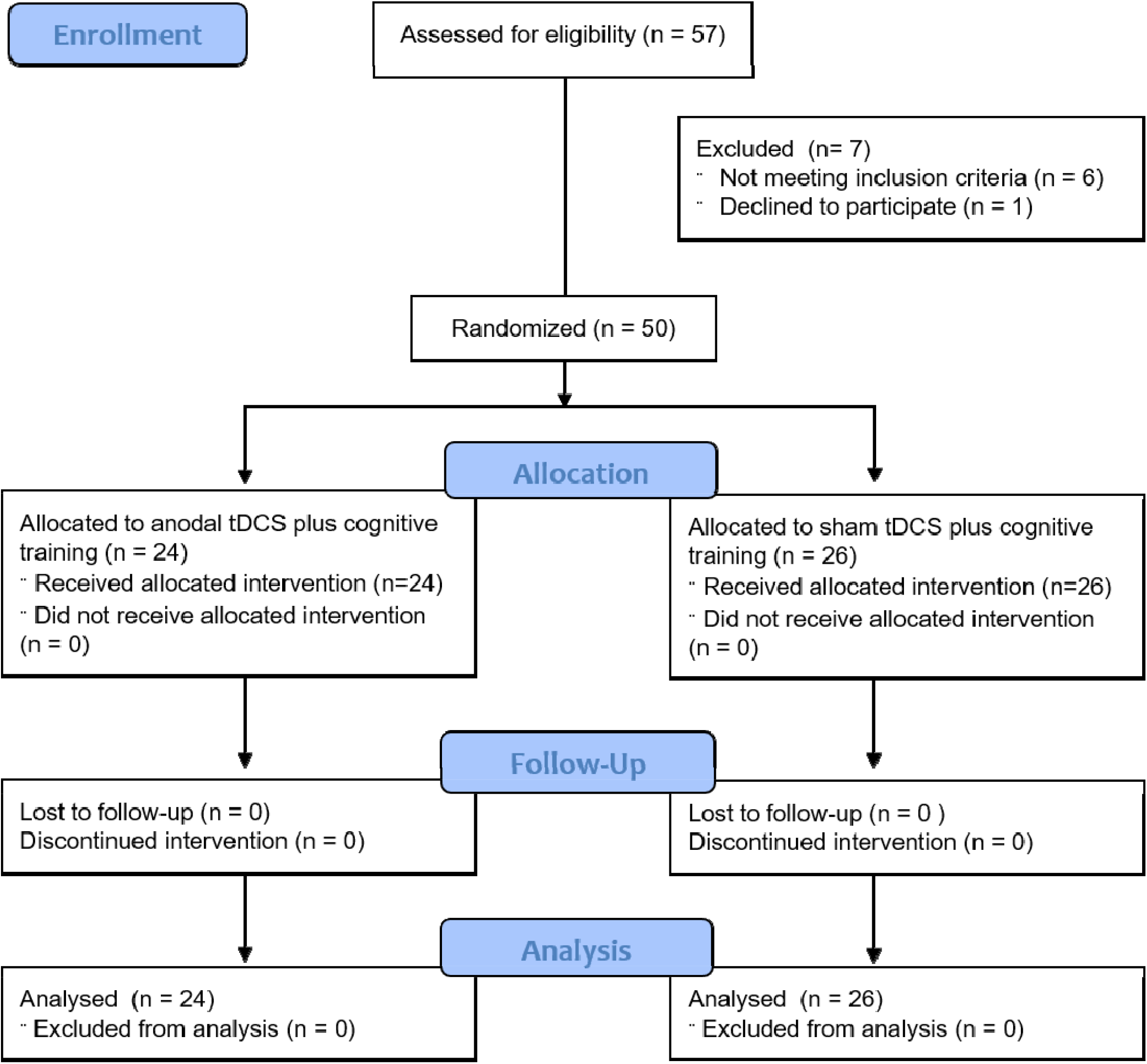
CONSORT flow diagram [56] of this RCT from enrolment, intervention allocation, follow-up, and analysis

Baseline assessment was scheduled at least two weeks after medication titration. Thirty-two participants received stable ADHD medication (non-psychostimulants: 2; psychostimulants: 30; between 21 weeks and 10 years). To help avoid medication effects from masking tDCS effects, 14 of the 32 participants followed our optional request to abstain from psychostimulants at least 24 hours before each assessment time point. A further seven participants abstained for trial duration (i.e., baseline to posttreatment). One participant stopped taking medication 3 months before follow-up.

### Intervention

Over 15 consecutive weekdays, participants played four 5-minute CT games during 20-minutes of anodal or sham tDCS. CT was composed of four ACTIVATE™ [62] games (Monday, Wednesday, and Friday) or a Stop task followed by three ACTIVATE™ games (Tuesday and Thursday). ACTIVATE™ games were played on an iPad Air 2 10.2”, the Stop task on a Dell XPS 15.6 Inch QHD Touch Laptop (Figure 2).

**Figure 2:**
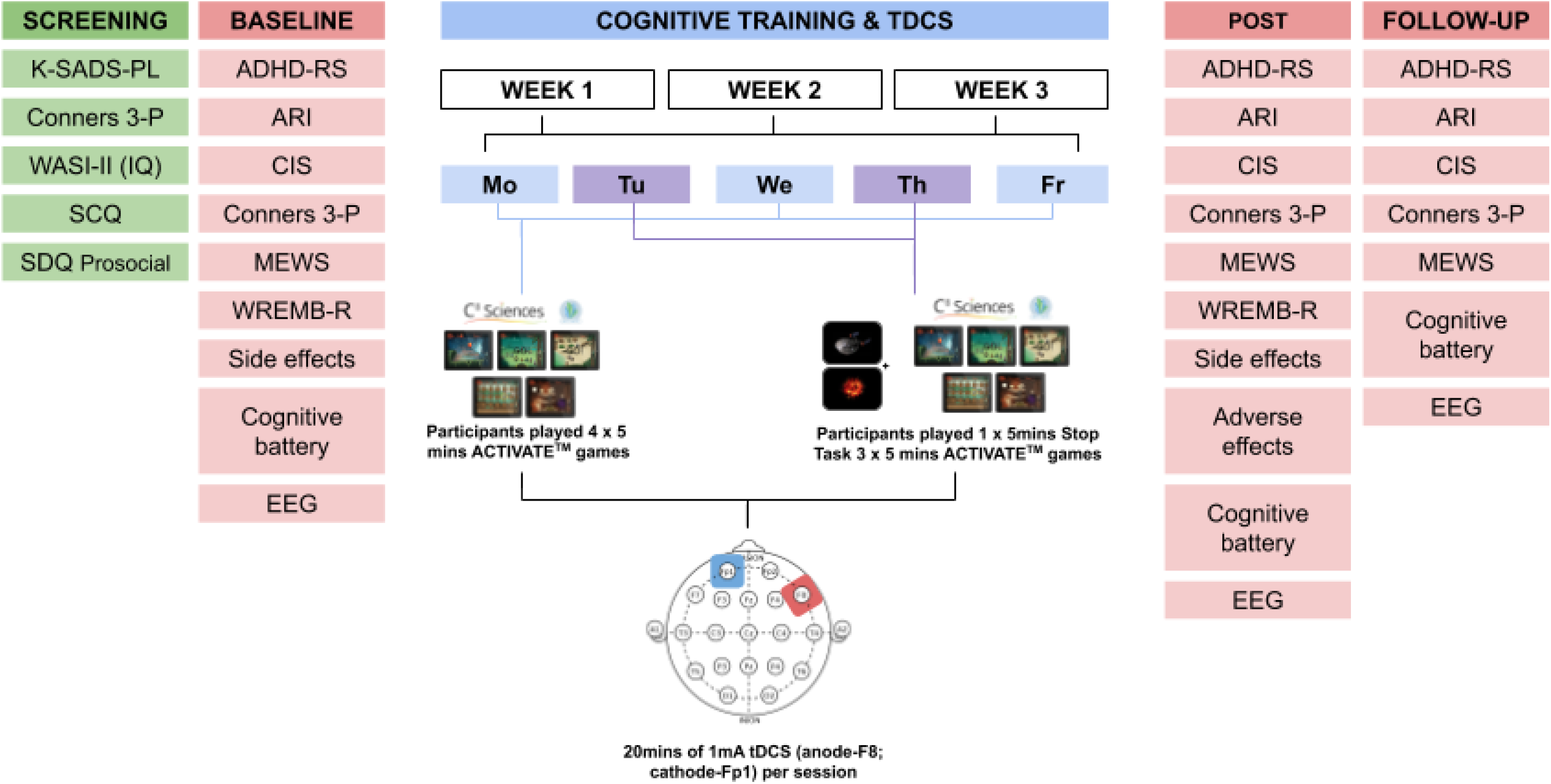
Schematic overview of the study design. ADHD-RS, Attention Deficit Hyperactivity Disorder-Rating Scale; ARI, Affective Reactivity Index; CIS, Columbia Impairment Scale; Conners 3-P, Conners’ 3^rd^ Edition Parent Rating; Cognitive battery, Maudsley Attention and Response Suppression task battery, vigilance, Wisconsin card sorting task, visual-spatial working memory, verbal fluency; K-SADS-PL, Kiddie-SADS-Present and Lifetime Version; MEWS, Mind Excessively Wandering Scale; SCQ, Social Communication Questionnaire (Lifetime), SDQ, Social Difficulties Questionnaire (prosocial scale only); WASI-II, Wechsler Abbreviated Scale of Intelligence, 2nd Edition; WREMB-R, Weekly Rating of Evening and Morning Behavior-Revised.

### CT games

The ACTIVATE™ programme trains multiple EF with engaging video games that increase in complexity as the player improves. Five of the six tasks were selected for this study to target ADHD-relevant EF, including selective and multiple simultaneous attention, inhibition and cognitive flexibility (i.e., *Magic Lens, Peter’s Printer Panic, Treasure Trunk*) and visual-spatial working memory (i.e., *Grub Ahoy, Monkey Trouble*). Before commencing each game, participants chose a game to play from three options; the ACTIVATE™ programme tracked choices and constrained future options so that multiple EF were twice as likely to be trained as working memory [62].

The training tracking Stop task trained motor response inhibition by requiring participants to cancel a prepotent motor response to a go signal that is followed shortly by an unexpected and rare (30% of trials) stop signal [63]. The delay between go and stop signals is dynamically adjusted to the participant performance, with better inhibition resulting in longer stop signal delays thus increasing the difficulty to withhold a response. Participants were encouraged to wait for the stop signal before responding to the go signal, thereby training inhibition and waiting behaviour/response delay (see Supplement).

### tDCS

Direct current was delivered by NovoStim (InnoSphere, Haifa, Israel) via a pair of 25 cm^2^ brush electrodes^1^ dipped in saline solution (0.45 mol). The anode was placed over rIFC (F8, according to the international EEG 10-20 system) [64–67], the cathode over the contralateral supraorbital area (Fp1), and both were held in place with a 10-20 EEG cap. For anodal tDCS, 1mA was administered for 20 minutes (current density: 0.04cm^2^; total charge: 0.8C/cm^2^) with a 30-second fade-in/fade-out. For sham stimulation, only the 30-second fade-in/fade-out was administered [68]. After each week, participants, parents, and tDCS administrators were asked to guess if participants had received anodal or sham tDCS.

### Primary Outcomes

#### Cognitive

The adult version of the Maudsley Attention and Response Suppression (MARS) task battery [63,69] measured motor response inhibition (GNG; dependent variable [DV]: probability of inhibition [PI]) and sustained attention (Continuous Performance Task [CPT]; DV: omission and commission errors) (see Supplement).

#### ADHD Symptoms

Parent-rated ADHD Rating Scale–IV (ADHD-RS) Home Version (DV: Total Scores) [70], a standard measure of treatment effects in DSM-IV ADHD symptoms.

### Secondary Outcomes

#### Cognitive Measures

MARS tasks [63,69] measured interference inhibition (Simon Task; DV: Simon reaction time effect) or time discrimination (Time Discrimination task; DV: percentage correct). The Mackworth Clock task [71–73] measured vigilance (DV: percentage omissions and commission errors), the Wisconsin Card Sorting Task [74] (WCST; DV: total and preservative errors) measured cognitive flexibility, and the C8 Sciences version of the NIH List Sorting Working Memory (WM) task (DV: total score) [76] measured visuo-spatial working memory. Phonetic and semantic fluency (DV: percentage of correct responses) [77] were measured to account for potential downregulation of lIFC mediated language production functions via interhemispheric inhibition.

#### Generic EF

Three intercorrelated generic, task-independent EF were measured, which included: 1) general speed of information processing (i.e. mean reaction times (MRT) for Go [GNG], Congruent [Simon], and Target [CPT] trials, weighted by number of trials) 2); intrasubject response variability (i.e., Coefficient of Variance [CV; SD of MRT divided by MRT] for Go [GNG], Congruent [Simon], and Target [CPT] trials); and 3) prematurity (i.e., premature responses to all trials in GNG, Simon and CPT) (see Supplement Table 1-5 and Figures 1-3).

#### ADHD symptoms and related impairments

Caregiver-rated Conners 3-P ADHD index [58] measured ADHD symptoms, Weekly Parent Ratings of Evening and Morning Behaviour-Revised scale (WREMB-R) [78] and the Columbia Impairment Scale-Parent version (CIS) [79] measured ADHD related difficulties and functional impairments, the child- and caregiver-rated Affective Reactivity Index (ARI) measured irritability [80], and the child-rated Mind Excessively Wandering Scale (MEWS) measured mind-wandering [81].

#### Safety, feasibility, and tolerability measures

Safety was measured with parent-rated questionnaires on side effects [82] and adverse events [83]. Participants rated their mood, wakefulness [84], and the tolerability of tDCS [85]. Parents and participants rated the overall impression of tDCS and CT using a rating scale from 0 (not at all/never) to 4 (definitely/always) designed specifically for this study.

#### EEG

EEG was measured at rest and during the GNG task at each assessment timepoint but will be reported in a separate publication.

#### Training outcome measures

The ACTIVATE™ DV was the highest game level reached each week averaged across three games that loaded on multiple EF (i.e., *Treasure Trunk, Magic Lens, Printer Panic*). The Stop task DV was PI on average for each week.

### Assessment Time Points

Primary and secondary outcome measures were collected at baseline, posttreatment, and six-month follow-up. Adverse events and overall impression of tDCS and CT were measured at posttreatment. Ratings of mood, wakefulness, and the tolerability of tDCS were measured in each stimulation session. Baseline and posttreatment sessions were scheduled up to three weeks before the first and after the last stimulation session, respectively.

### Statistical Methods

#### Confirmatory analysis

Repeated measures analysis of covariance (ANCOVA) tested group differences across posttreatment and follow-up, covarying for age in months, medication status (naïve, on-, or off-medication), and baseline value of outcomes where applicable. Sensitivity analyses were also conducted to remove statistical outliers on cognitive tasks, participants who changed medication between posttreatment and follow-up, or participants whose treatment spanned over four instead of three weeks (see Supplement Table 6-8). The alpha level was set to 0.05, and False Discovery Rate (FDR) correction with Benjamini-Hochberg p-value adjustment [86] was applied to control for multiple testing. Only results based on adjusted *p*-values are reported, see Tables 3-4 for unadjusted p-values. Analyses were conducted with IBM SPSS Statistics 26 (IBM Corp., Armonk, N.Y., USA).

**Table 1:**
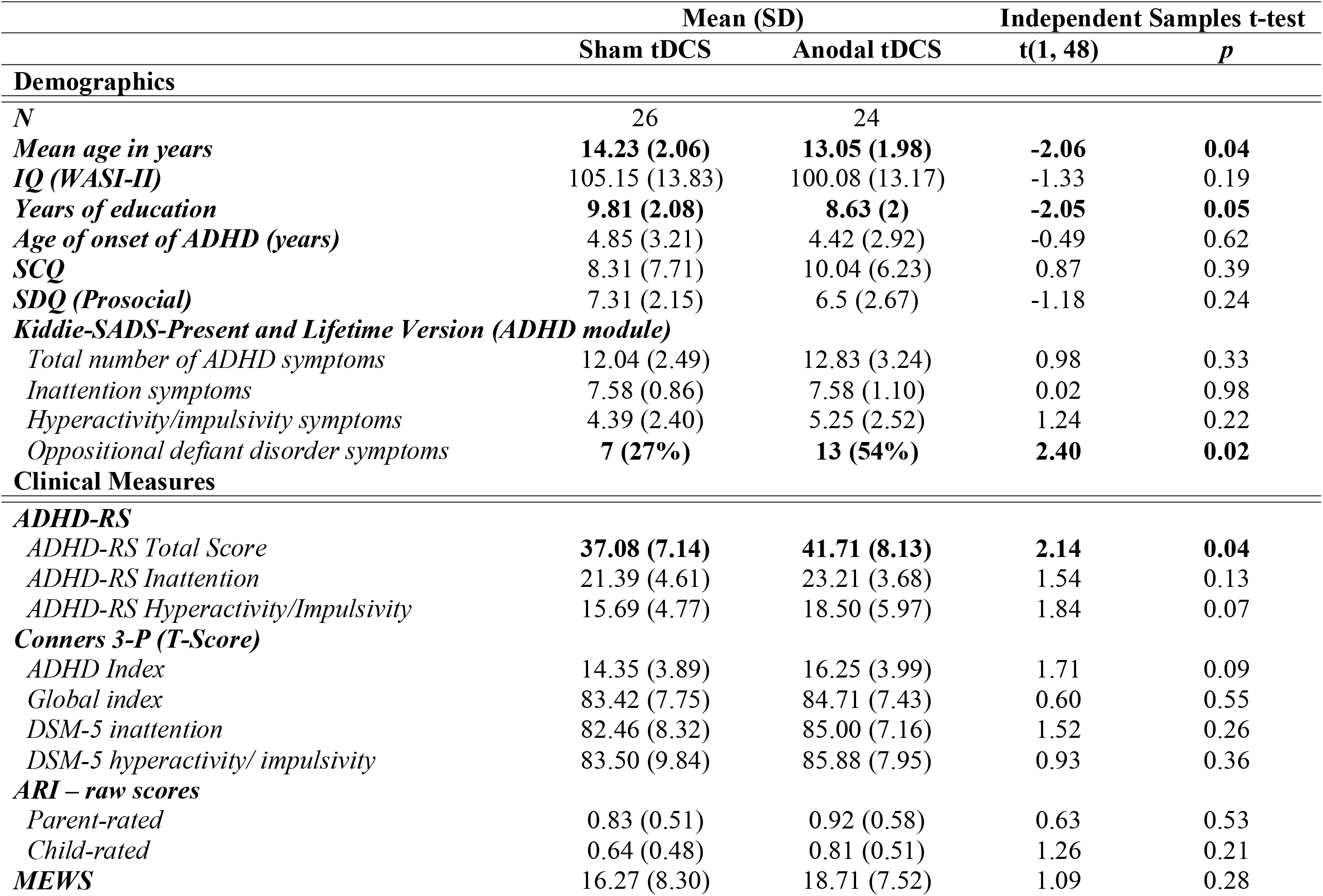
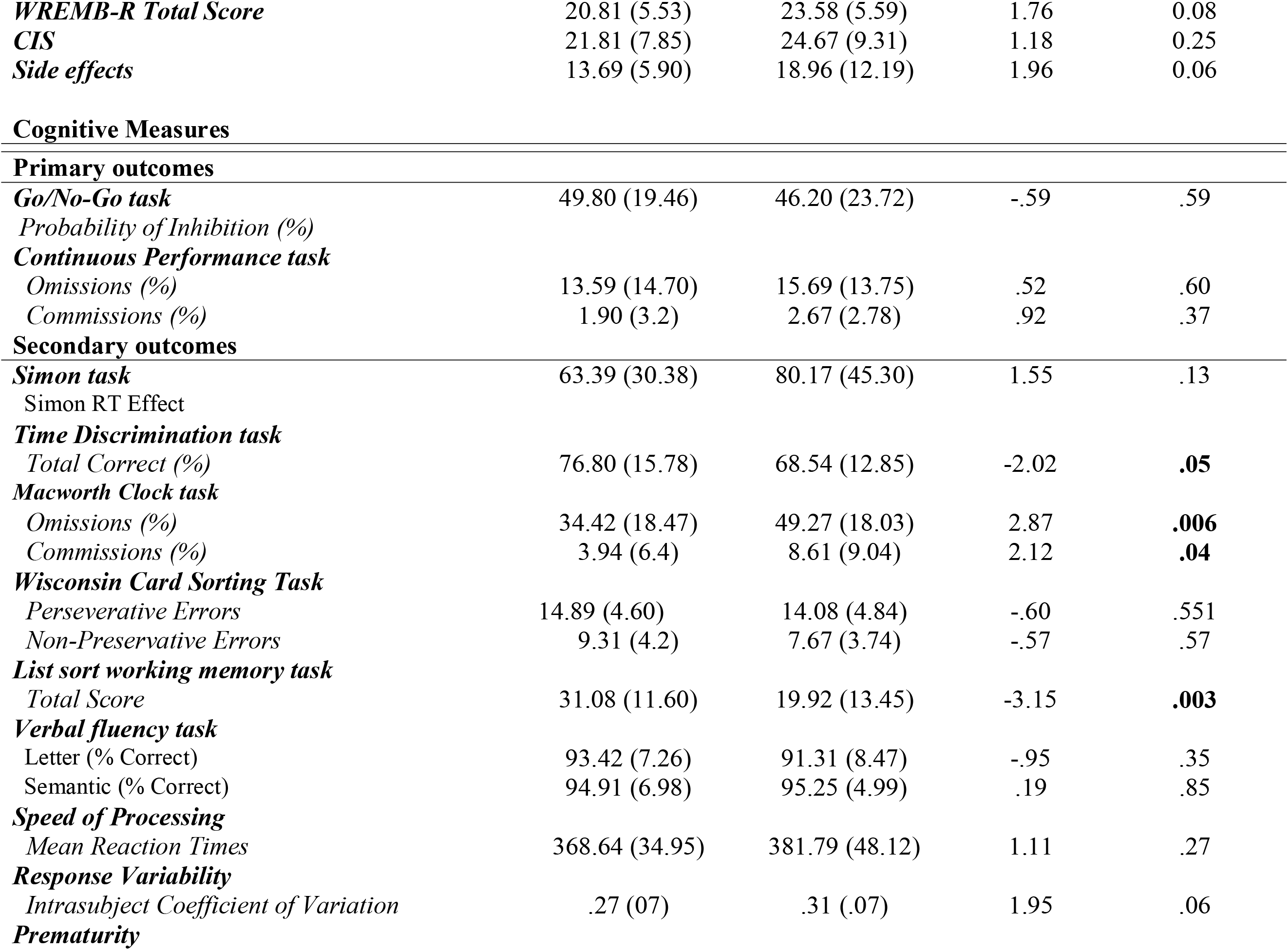
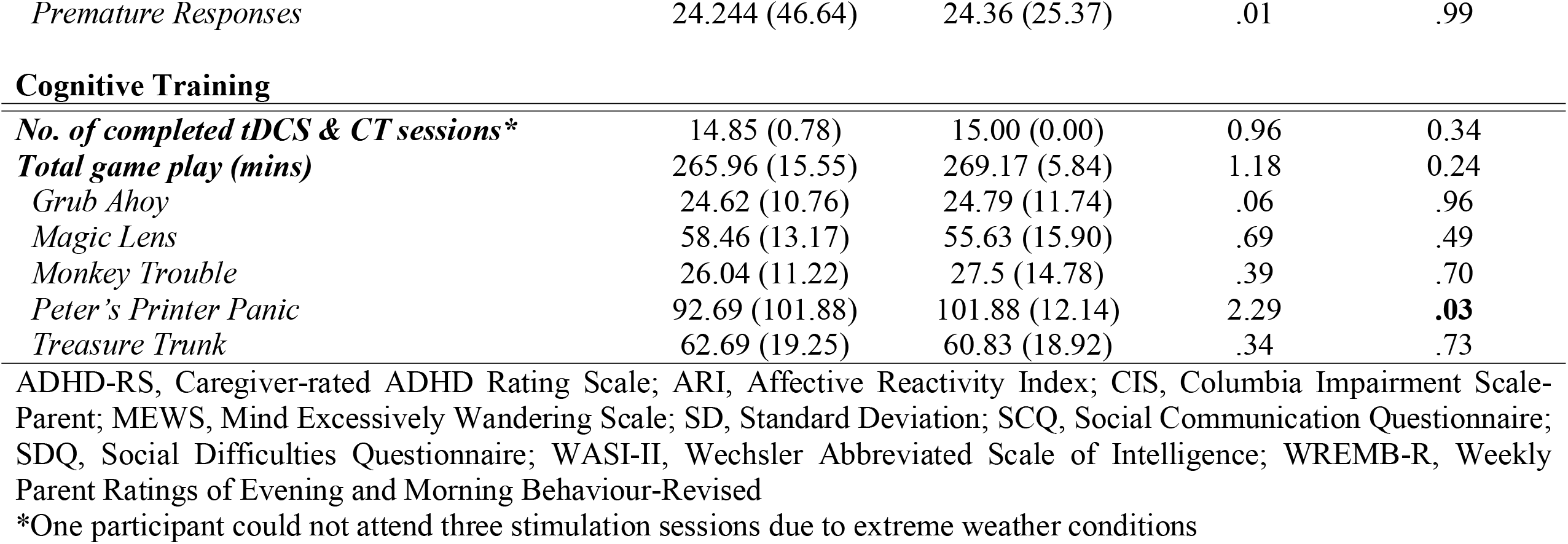
Baseline demographic, clinical, and cognitive measures; the number of tDCS and CT sessions; and the time spent playing each CT game in the sham and anodal tDCS groups

|  | Mean (SD) |  | Independent Samples t-test |  |
| --- | --- | --- | --- | --- |
|  | Sham tDCS | Anodal tDCS | t(1, 48) | p |
| **Demographics** |  |  |  |  |
| *N* | 26 | 24 |  |  |
| *Mean age in years* | **14.23 (2.06)** | **13.05 (1.98)** | **-2.06** | **0.04** |
| *IQ (WASI-II)* | 105.15 (13.83) | 100.08 (13.17) | -1.33 | 0.19 |
| *Years of education* | **9.81 (2.08)** | **8.63 (2)** | **-2.05** | **0.05** |
| *Age of onset of ADHD (years)* | 4.85 (3.21) | 4.42 (2.92) | -0.49 | 0.62 |
| *SCQ* | 8.31 (7.71) | 10.04 (6.23) | 0.87 | 0.39 |
| *SDQ (Prosocial)* | 7.31 (2.15) | 6.5 (2.67) | -1.18 | 0.24 |
| *Kiddie-SADS-Present and Lifetime Version (ADHD module)* |  |  |  |  |
| *Total number of ADHD symptoms* | 12.04 (2.49) | 12.83 (3.24) | 0.98 | 0.33 |
| *Inattention symptoms* | 7.58 (0.86) | 7.58 (1.10) | 0.02 | 0.98 |
| *Hyperactivity/impulsivity symptoms* | 4.39 (2.40) | 5.25 (2.52) | 1.24 | 0.22 |
| *Oppositional defiant disorder symptoms* | **7 (27%)** | **13 (54%)** | **2.40** | **0.02** |
| **Clinical Measures** |  |  |  |  |
| **ADHD-RS** |  |  |  |  |
| *ADHD-RS Total Score* | **37.08 (7.14)** | **41.71 (8.13)** | **2.14** | **0.04** |
| *ADHD-RS Inattention* | 21.39 (4.61) | 23.21 (3.68) | 1.54 | 0.13 |
| *ADHD-RS Hyperactivity/Impulsivity* | 15.69 (4.77) | 18.50 (5.97) | 1.84 | 0.07 |
| **Conners 3-P (T-Score)** |  |  |  |  |
| *ADHD Index* | 14.35 (3.89) | 16.25 (3.99) | 1.71 | 0.09 |
| *Global index* | 83.42 (7.75) | 84.71 (7.43) | 0.60 | 0.55 |
| *DSM-5 inattention* | 82.46 (8.32) | 85.00 (7.16) | 1.52 | 0.26 |
| *DSM-5 hyperactivity/ impulsivity* | 83.50 (9.84) | 85.88 (7.95) | 0.93 | 0.36 |
| **ARI – raw scores** |  |  |  |  |
| *Parent-rated* | 0.83 (0.51) | 0.92 (0.58) | 0.63 | 0.53 |
| *Child-rated* | 0.64 (0.48) | 0.81 (0.51) | 1.26 | 0.21 |
| **MEWS** | 16.27 (8.30) | 18.71 (7.52) | 1.09 | 0.28 |
| **WREMB-R Total Score** | 20.81 (5.53) | 23.58 (5.59) | 1.76 | 0.08 |
| **CIS** | 21.81 (7.85) | 24.67 (9.31) | 1.18 | 0.25 |
| **Side effects** | 13.69 (5.90) | 18.96 (12.19) | 1.96 | 0.06 |
| Cognitive Measures |  |  |  |  |
| **Primary outcomes** |  |  |  |  |
| **Go/No-Go task** | 49.80 (19.46) | 46.20 (23.72) | -.59 | .59 |
| *Probability of Inhibition (%)* |  |  |  |  |
| **Continuous Performance task** |  |  |  |  |
| *Omissions (%)* | 13.59 (14.70) | 15.69 (13.75) | .52 | .60 |
| *Commissions (%)* | 1.90 (3.2) | 2.67 (2.78) | .92 | .37 |
| **Secondary outcomes** |  |  |  |  |
| **Simon task** | 63.39 (30.38) | 80.17 (45.30) | 1.55 | .13 |
| Simon RT Effect |  |  |  |  |
| **Time Discrimination task** |  |  |  |  |
| *Total Correct (%)* | 76.80 (15.78) | 68.54 (12.85) | -2.02 | **.05** |
| **Macworth Clock task** |  |  |  |  |
| *Omissions (%)* | 34.42 (18.47) | 49.27 (18.03) | 2.87 | **.006** |
| *Commissions (%)* | 3.94 (6.4) | 8.61 (9.04) | 2.12 | **.04** |
| **Wisconsin Card Sorting Task** |  |  |  |  |
| *Perseverative Errors* | 14.89 (4.60) | 14.08 (4.84) | -.60 | .551 |
| *Non-Preservative Errors* | 9.31 (4.2) | 7.67 (3.74) | -.57 | .57 |
| **List sort working memory task** |  |  |  |  |
| *Total Score* | 31.08 (11.60) | 19.92 (13.45) | -3.15 | **.003** |
| **Verbal fluency task** |  |  |  |  |
| Letter (% Correct) | 93.42 (7.26) | 91.31 (8.47) | -.95 | .35 |
| Semantic (% Correct) | 94.91 (6.98) | 95.25 (4.99) | .19 | .85 |
| **Speed of Processing** |  |  |  |  |
| *Mean Reaction Times* | 368.64 (34.95) | 381.79 (48.12) | 1.11 | .27 |
| **Response Variability** |  |  |  |  |
| *Intrasubject Coefficient of Variation* | .27 (.07) | .31 (.07) | 1.95 | .06 |
| **Prematurity** |  |  |  |  |
| *Premature Responses* | 24.244 (46.64) | 24.36 (25.37) | .01 | .99 |
| **Cognitive Training** |  |  |  |  |
| *No. of completed tDCS & CT sessions\** | 14.85 (0.78) | 15.00 (0.00) | 0.96 | 0.34 |
| *Total game play (mins)* | 265.96 (15.55) | 269.17 (5.84) | 1.18 | 0.24 |
| *Grub Ahoy* | 24.62 (10.76) | 24.79 (11.74) | .06 | .96 |
| *Magic Lens* | 58.46 (13.17) | 55.63 (15.90) | .69 | .49 |
| *Monkey Trouble* | 26.04 (11.22) | 27.5 (14.78) | .39 | .70 |
| *Peter's Printer Panic* | 92.69 (101.88) | 101.88 (12.14) | 2.29 | **.03** |
| *Treasure Trunk* | 62.69 (19.25) | 60.83 (18.92) | .34 | .73 |

ADHD-RS, Caregiver-rated ADHD Rating Scale; ARI, Affective Reactivity Index; CIS, Columbia Impairment Scale-Parent; MEWS, Mind Excessively Wandering Scale; SD, Standard Deviation; SCQ, Social Communication Questionnaire; SDQ, Social Difficulties Questionnaire; WASI-II, Wechsler Abbreviated Scale of Intelligence; WREMB-R, Weekly Parent Ratings of Evening and Morning Behaviour-Revised
\*One participant could not attend three stimulation sessions due to extreme weather conditions

**Table 2.** Medication status in the sham and anodal tDCS groups

| ADHD Medication Status | N (%) |  | Pearson Chi Square |  |
| --- | --- | --- | --- | --- |
| | Sham tDCS<br>(n = 26) | Anodal tDCS<br>(n = 24) | $\chi^2$ (3) | p |
| *Medication-naïve* | 8 (31%) | 10 (42%) | 6.28 | 0.10 |
| *On medication* | 3 (27%) | 8 (33%) |  |  |
| *On-medication except for assessments* | 10 (39%) | 4 (17%) |  |  |
| *Off medication* | 5 (19%) | 2 (8%) |  |  |
| ADHD Medication Type | | | $\chi^2$ (5) | p |
| *Atomoxetine* | 0 | 1 (4%) | 7.54 | .18 |
| *Dexamfetamine* | 0 | 1 (4%) |  |  |
| *Guanfacine* | 0 | 2 (8%) |  |  |
| *Lisdexamfetamine* | 2 (8%) | 0 |  |  |
| *Methylphenidate* | 16 (62%) | 10 (42%) |  |  |
| *Medication-naïve* | 8 (31%) | 10 (42%) |  |  |

**Table 3.** Summary of adjusted average performance on primary cognitive and clinical outcome measures after sham and anodal tDCS combined with CT. Benjamini-Hochberg adjusted p-values given parentheses.

| Primary Outcomes | Posttreatment |  | Follow-up |  | ANCOVA |  |  |  |  |  |
| --- | --- | --- | --- | --- | --- | --- | --- | --- | --- | --- |
|  | Sham tDCS | Anodal tDCS | Sham tDCS | Anodal tDCS | Time |  | Group |  | Time by Group |  |
|  | Adjusted Mean (SD)* |  |  |  | F (1,44) | p † | F (1,44) | p † | F (1,44) | p † |
| Cognitive | N=26 | N=24 | N=26 | N=24 |  |  |  |  |  |  |
| Go/No-Go task |  |  |  |  |  |  |  |  |  |  |
| PI (%) | 54.18 (14.18) | 55.48 (14.23) | 56.95 (16.96) | 58.41 (17.02) | .01 | .93 (.93) | .12 | .73 (.73) | .001 | .97 (.97) |
| Continuous Performance task |  |  |  |  |  |  |  |  |  |  |
| Omission (%) | 9.88 (8.44) | 14.36 (8.48) | 8.31 (8.22) | 11.70 (8.25) | .32 | .58 (1.00) | 3.87 | .06 (.09) | □.15 | .71 (1.00) |
| Commission (%) | 1.04 (1.65) | 2.16 (1.66) | 0.97 (1.14) | 1.26 (1.14) | .08 | .79 (1.00) | 4.96 | .03 (.09) | 2.67 | .14 (.42) |
| Clinical |  |  |  |  |  |  |  |  |  |  |
| ADHD-RS‡ |  |  |  |  |  |  |  |  |  |  |
| Total Score | 25.55 (8.91) | 32.48 (8.96) | 31.58 (9.51) | 27.91 (9.54) | 1.28 | .26 | .57 | .46 | 10.58 | .002 |
| Inattention | 14.61 (5.04) | 18.01 (5.05) | 16.87 (5.35) | 16.10 (5.34) | .04 | .84 | 1.41 | .24 | 4.02 | .051 |
| Hyperactivity/Impulsivity | 11.30 (4.95) | 14.10 (4.95) | 14.62 (5.00) | 11.91 (5.00) | .01 | .91 | .001 | .97 | 13.08 | .001 |

ADHD-RS, ADHD Rating Scale; PI, Probability of Inhibition; SD, Standard Deviation
\*Adjusted values as predicted by the repeated-measures ANCOVA testing group differences at posttreatment and follow-up, adjusting for baseline, age at entry and medication status (naïve, off-medication, on-medication).
†Benjamini-Hochberg adjustment was applied to p-values for time, group, and time by group interaction effect separately and was applied separately to primary cognitive, secondary cognitive, and secondary clinical outcome measures separately.
‡Benjamini-Hochberg adjustment was not applied to these measures

**Table 4.** Summary of adjusted average performance on secondary cognitive and clinical outcome measures after sham and anodal tDCS combined with CT. Benjamini-Hochberg adjusted p-values given parentheses.

| Secondary Outcomes | Posttreatment |  | Follow-up |  | ANCOVA |  |  |  |  |  |
| --- | --- | --- | --- | --- | --- | --- | --- | --- | --- | --- |
|  | Sham tDCS | Anodal tDCS | Sham tDCS | Anodal tDCS | Time |  | Group |  | Time by Group |  |
|  | *Adjusted Mean (SD)\** |  |  |  | *F (1,44)* | *p* | *F (1,44)* | *p* | *F (1,44)* | *p* |
| Cognitive | *N=26* | *N=24* | *N=26* | *N=24* |  |  |  |  |  |  |
| *Simon task* |  |  |  |  |  |  |  |  |  |  |
| Simon RT Effect | 45.78 (32.03) | 59.91 (32.15) | 49.92 (25.54) | 58.86 (25.64) | .11 | .74 (.89) | 2.52 | .12 (.36) | .32 | .58 (1.00) |
| *Time Discrimination task* |  |  |  |  |  |  |  |  |  |  |
| Total Correct (%) | 73.05 (12.31) | 65.38 (16.17) | 78.34 (11.78) | 70.93 (15.47) | 2.21 | .14 (.84) | 2.48 | .11 (.45) | .003 | .96 (1.00) |
| *Macworth Clock task* |  |  |  |  |  |  |  |  |  |  |
| Commissions (%) | 4.50 (4.57) | 4.91 (4.60) | 5.09 (8.09) | 4.80 (8.12) | 2.42 | .13 (1.00) | .001 | .93 (.93) | .25 | .62 (.93) |
| Omissions (%) | 34.26 (15.17) | 38.82 (15.25) | 27.24 (11.78) | 34.14 (11.85) | 1.79 | .19 (.46) | 2.89 | .10 (.60) | .26 | .61 (1.00) |
| *Wisconsin Card Sorting Task (errors)* |  |  |  |  |  |  |  |  |  |  |
| Non Perseverative | 7.11 (4.89) | 8.9 (4.89) | 7.11 (3.57) | 7.59 (3.72) | 1.83 | .18 (.54) | 1.21 | .28 (.48) | .88 | .35 (1.00) |
| Preservative | 12.20 (4.64) | 13.53 (4.65) | 13.39 (5.87) | 13.32 (5.89) | .503 | .48 (.82) | .27 | .61 (.73) | .51 | .48 (1.00) |
| *List sort working memory task* |  |  |  |  |  |  |  |  |  |  |
| Total Score | 27.57 (14.20) | 28.35 (14.65) | 31.27 (16.65) | 31.79 (17.18) | .14 | .71 (.95) | .03 | .86 (.94) | .002 | .97 (.97) |
| *Verbal Fluency task* |  |  |  |  |  |  |  |  |  |  |
| Letter % Corr | 96.92 (3.97) | 94.28 (3.98) | 97.13 (4.08) | 96.93 (4.09) | .27 | .60 (.90) | 2.47 | .12 (.29) | 2.52 | .12 (.72) |
| Semantic % Corr | 96.24 (3.69) | 97.08 (3.70) | 98.08 (5.32) | 94.89 (5.33) | .12 | .74 (.81) | 1.22 | .28 (.56) | 5.89 | .02 (.24) |
| *Speed of Processing* |  |  |  |  |  |  |  |  |  |  |
| MRT | 382.86 (31.02) | 393.21 (31.12) | 361.39 (31.32) | 367.46 (31.42) | 1.97 | .17 (.68) | 1.16 | .29 (.43) | .17 | .68 (.91) |
| *Response Variability* |  |  |  |  |  |  |  |  |  |  |
| ICV | .27 (.00) | .28 (.00) | .25 (.00) | .26 (.00) | 1.09 | .30 (.60) | .78 | .38 (.51) | .06 | .81 (.97) |
| *Prematurity* |  |  |  |  |  |  |  |  |  |  |
| Premature Resp. | 13.55 (4.10) | 24.99 (4.28) | 7.78 (1.85) | 8.39 (1.93) | .10 | .75 (.75) | 3.32 | .08 (.96) | 2.46 | .12 (.48) |
| **Clinical** |  |  |  |  |  |  |  |  |  |  |
| *Conners 3-P* | 8.01 (4.41) | 12.53 (4.42) | 11.72(4.94) | 11.22 (4.95) | .001 | .97 (.97) | 2.88 | .10 (.30) | 13.73 | .001 (.004) |
| **ADHD Index** |  |  |  |  |  |  |  |  |  |  |
| **ARI** |  |  |  |  |  |  |  |  |  |  |
| Parent | 0.72 (.47) | 0.79 (.44) | 0.67 (.45) | 0.49 (.42) | .27 | .61 (.81) | 2.77 | .10 (.20) | 2.89 | .20 (.10) |
| Child | 0.56 (.29) | 0.55 (.5) | 0.61 (.40) | 0.60 (.4) | .702 | .41 (.82) | .02 | .89 (.89) | .001 | .99 (.99) |
| **MEWS** | 16.16 (6.22) | 16.04 (6.23) | 18.08 (6.92) | 14.87 (6.94) | 5.01 | **.03** (.12) | 1.08 | .31 (.37) | 2.18 | .20 (.15) |
| **WREMB-R** | 15.21 (6.09) | 18.28 (6.10) | n/t | n/t | n/a | n/a | 2.96 | .09 (.54) | n/a | n/a |
| **CIS** | 16.75 (8.55) | 19.82 (8.56) | n/t | n/t | n/a | n/a | 1.52 | .22 (.33) | n/a | n/a |
| **Safety‡** |  |  |  |  |  |  |  |  |  |  |
| Side Effects | 11.92 (7.02) | 14.59 (7.03) | n/t | n/t | n/a | n/a | 1.68 | .20 | n/a | n/a |
| Adverse Effects | 15.04 (2.20) | 16.33 (2.20) | n/t | n/t | n/a | n/a | **4.09** | **.05** | n/a | n/a |

ARI, Affective Reactivity Index; CIS, Columbia Impairment Scale-Parent; Corr, Correct; ICV, intrasubject coefficient of variance; MEWS, Mind Excessively Wandering Scale; MRT, Mean Reaction Times; Resp, responses; SD, Standard Deviation; WREMB-R, Weekly Parent Ratings of Evening and Morning Behaviour-Revised
\*Adjusted values as predicted by the repeated-measures ANCOVA testing group differences at posttreatment and follow-up, adjusting for baseline, age at entry and medication status (naïve, off-medication, on-medication).
†Benjamini-Hochberg adjustment was applied to p-values for time, group, and time by group interaction effects separately and was applied separately to primary cognitive, secondary cognitive, and secondary clinical outcome measures separately.
‡Benjamini-Hochberg adjustment was not applied to these measures

#### Medication status

Medication status was coded as a categorical covariate with three groups: medication-naïve, on-medication, and off-medication. Participants who abstained for the treatment trial period were coded as off-medication, participants who abstained at assessment time points only were coded as off-medication for cognitive outcomes and on-medication for clinical outcomes (which can cover three weeks).

#### Missing data & statistical outliers

In primary and secondary outcomes, only posttreatment WM task data for one participant were missing. Missing stop signal task data (1.33%) were random and replaced by group averages.

## RESULTS

### Baseline demographics

Compared to sham tDCS, the anodal tDCS group was significantly younger, had fewer years in education, higher ADHD-RS Total scores and ODD symptoms, and worse performance on the Macworth Clock, Time Discrimination, and list sort working memory tasks and during CT spent significantly more time playing the *Peter’s Printer Panic* game (Tables 1 and 2).

### Primary outcome measures

#### Cognitive

There were no significant effects on primary cognitive outcome measures after adjusting for multiple testing (see Table 3 and Figure 3).

**Figure 3.**
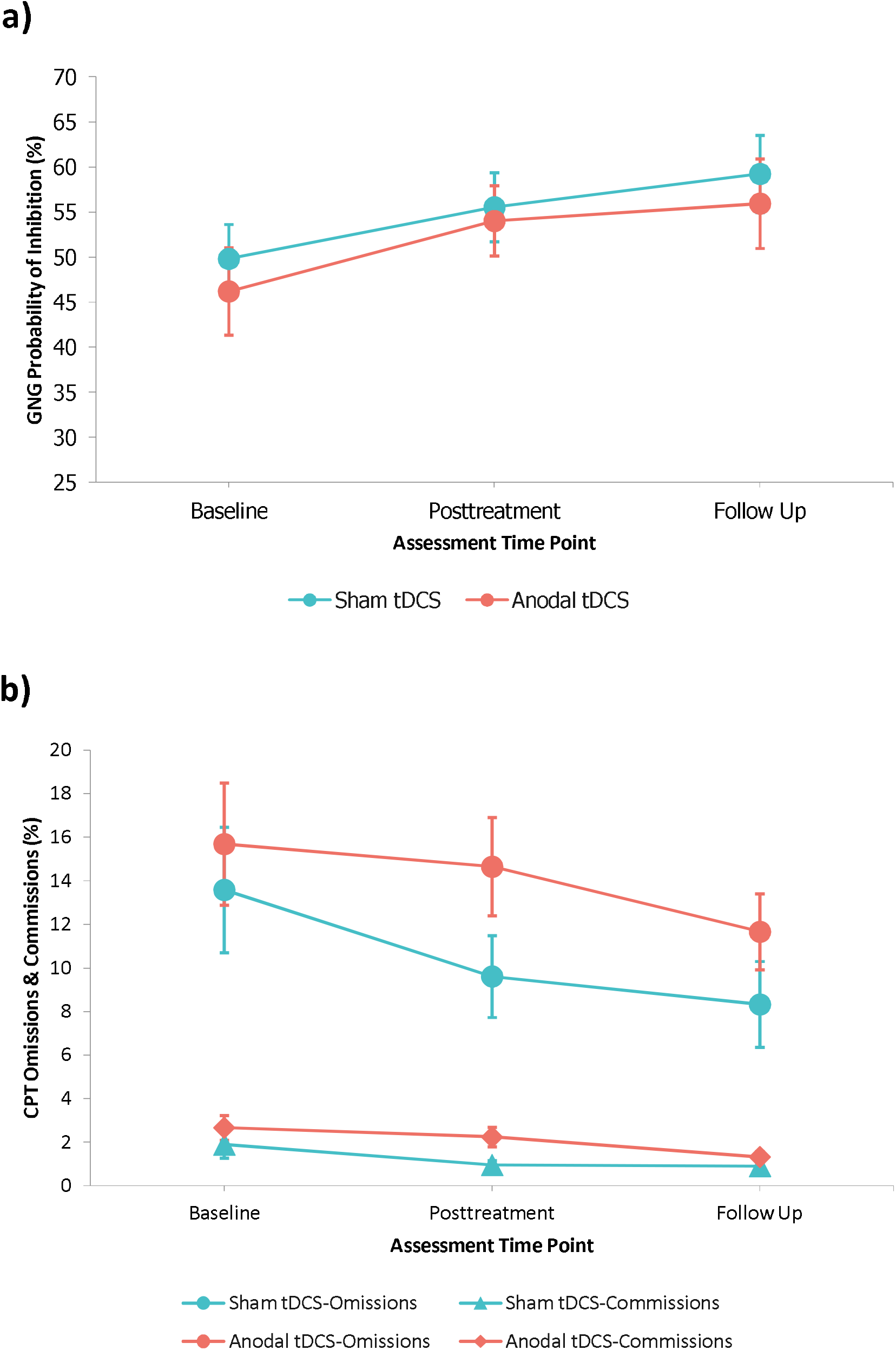
Raw averages for a) GNG Probability of Inhibition (%) and b) CPT Omissions at baseline, posttreatment, and follow-up for sham and anodal tDCS groups (error bars: standard error).

#### ADHD symptoms

There was only a significant time by group interaction (*F*_1,44_=10.58, *p*=.002, ηp^2^=.19); simple effects analysis showed that the anodal versus sham tDCS group had higher scores at posttreatment but not at follow-up (posttreatment: *p*=.011 [95% CI: 1.65, 12.21]; follow-up: *p*=.20 [95% CI: −9.30, 1.97]). To determine what drove this effect, exploratory simple effects analysis of subscales showed that the anodal versus sham tDCS group had higher scores at posttreatment on the Inattention (*p*=0.03) and Hyperactivity-Impulsivity subscales (*p*=.06), with the latter being lower for active vs. sham tDCS at follow-up (*p*=0.07) (Table 3 and Figure 4).

**Figure 4.**
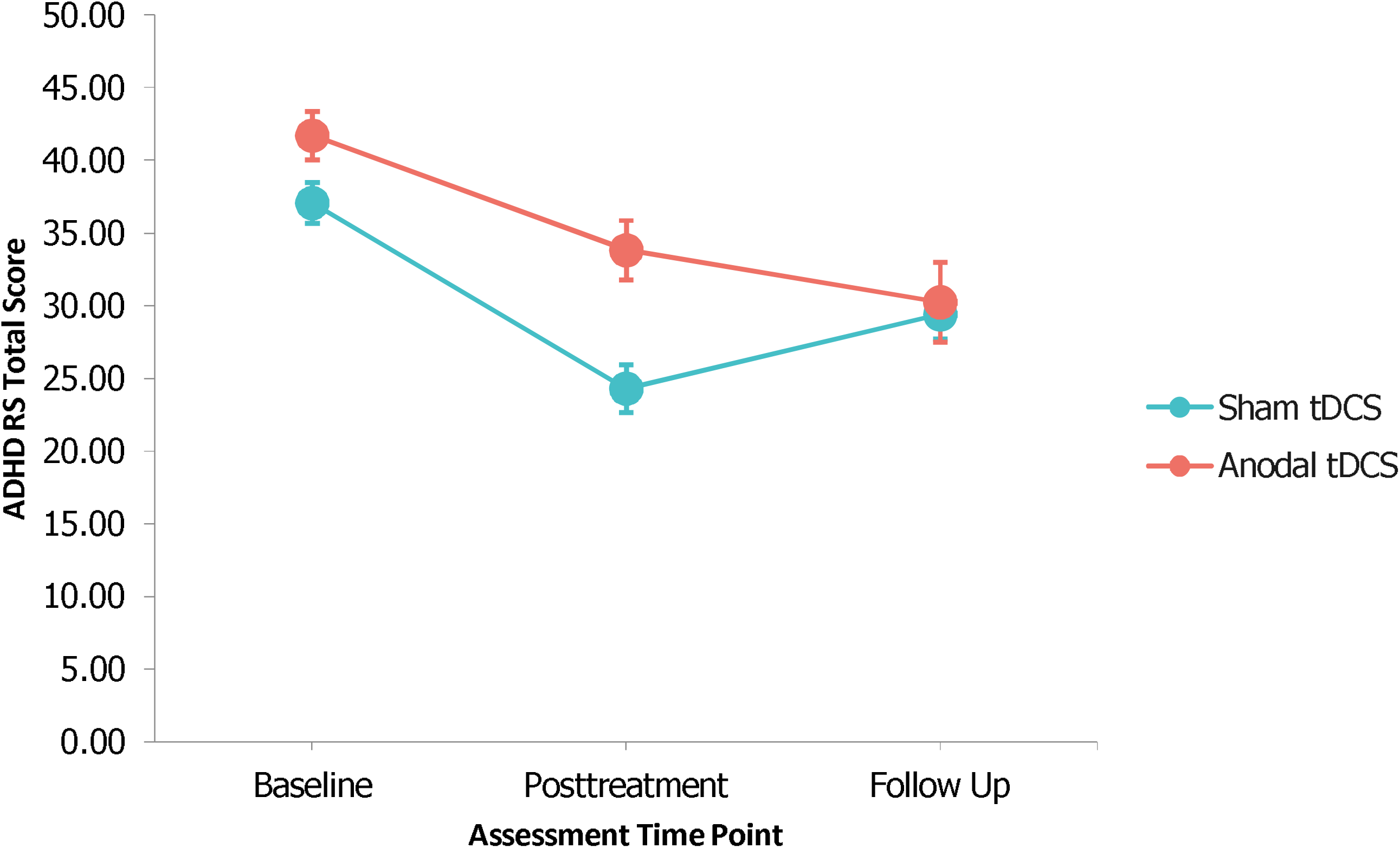
Raw average ADHD-RS Total Scores at baseline, posttreatment, and follow-up for sham tDCS and anodal tDCS groups (error bars: standard error).

### Secondary outcomes measures

#### Cognitive

The were no significant effects on secondary cognitive outcome measures after adjusting for multiple testing (See Table 4).

#### ADHD symptoms and related impairments

There was only a significant time by group interaction for Conners 3-P ADHD Index (*F*_1,44_=13.726, *p*=.004, ηp^2^=.238). Simple effects analysis showed significantly higher scores in the anodal versus sham tDCS group at posttreatment only (*p*=.001 [95%CI: 1.92, −7.11]; follow-up: *p*=.73 [95%CI: −3.39, 2.41]) (Table 4).

### Exploratory analyses

Separate repeated measures ANOVAs showed an effect of time (baseline versus posttreatment or follow-up) across groups. In cognitive measures, both groups improved in GNG task, Simon task, Letter Fluency (baseline versus posttreatment or follow-up);, Speed of Processing (baseline versus posttreatment only); and CPT % Omissions and Commissions, Mackworth Clock % Omissions, WM, Response Variability, and Prematurity (baseline versus follow-up only) (see Table 9 in Supplement). In clinical measures, both groups improved in ADHD-RS and Conners 3-P Index (baseline versus posttreatment or follow-up); and in ARI Parent and Child, WREMB-R, and CIS (baseline versus posttreatment only).

Group differences in CT performance across the three weeks were explored with repeated□measures ANCOVAs covarying for baseline, age in years, medication status (naïve, on-, or off-medication) and total time spent playing each game. There were significant effects in ACTIVATE™ or Stop task after adjusting for multiple testing (see Table 10 in Supplement).

We also explored if outcome changes that showed a significant time effect across both groups from baseline to posttreatment or follow-up were correlated with changes in CT performance scores (week 3 minus week 1). No correlations were significant (Table 11 in Supplement).

Given that age was not matched between groups, we conducted an post-hoc moderation analysis [87], predicting a change in ADHD-RS (baseline minus posttreatment) in a regression analysis by stimulation group, age, and a stimulation by age interaction. While the stimulation by age interaction was not significant (β=0.2, *SE*=.12, *t*(46)=1.7, *p*=.096), the simple effects showed a significant reduced improvement for anodal tDCS versus sham in older participants (1SD above mean age; β=9.53, *SE*=4.26, *t*(46)=2.24, *p*=.03), and an opposite but not significant pattern in younger children (1SD below mean age; β=-.6, *SE*=4.11, *t*(46)=-.15, *p*=.88), indicating that older participants benefitted clinically less from anodal versus sham tDCS.

### Safety, feasibility, tolerability, and blinding integrity

There were no significant group differences in ratings of mood, wakefulness, overall impression of tDCS and CT (Tables 12 and 13 in Supplement), and side effects (Table 4). Adverse effects were significantly higher in the anodal versus sham tDCS group at posttreatment (*F*_*1,44*_=4.09, *p=*.05, ηp^2^=.08), driven mainly by higher parent-ratings for the items “he seems more grumpy and irritable”, “has little appetite”, and “has more problems falling asleep” (Table 4). Tolerability ratings showed that stimulation was well tolerated, with only significantly higher reports of burning sensation during anodal than sham tDCS (see Table 14 in Supplement). Group assignment guesses did not exceed chance level for experimenters (χ^2^[1]=3.9, *p*=.28) and participants (χ^2^[1]=1.85, *p*=.17), with only parent guesses for anodal tDCS reaching borderline significance (χ^2^(1)=3.57, *p*=.06), thus blinding was overall successful.

## DISCUSSION

This double-blind, sham-controlled RCT showed that 15 sessions of anodal compared to sham tDCS over rIFC combined with CT in 50 boys with ADHD showed no improvement in ADHD symptoms or cognitive performance. Although both groups improved in clinical and cognitive measures over time, anodal relative to sham tDCS was associated with higher primary (ADHD-RS) and secondary (Conners 3-P ADHD Index) clinical outcome measures. Side effects did not differ, but at posttreatment, higher adverse effects relating to mood, sleep and appetite were reported following anodal compared to sham tDCS.

The lack of an observable clinical or cognitive effect extend previous meta-analytic evidence of no significant cognitive effects and limited evidence of clinical effects in ADHD with 1-5 sessions of predominantly left dlPFC anodal tDCS [19]. These findings are unexpected given that rIFC underactivation is consistently associated with poor cognitive control, attention and clinical symptoms in ADHD [3,7–9]. While the findings of no clinical effect of tDCS of rIFC are novel, the negative effects on cognition extend evidence from prior 1 or 5 session sham-controlled tDCS studies stimulating rIFC in ADHD which showed no or moderate effects (see introduction) [51–53].

Findings are furthermore unexpected given evidence of a synergistic effect of combined CT and tDCS on improving cognition [32,34,88]. Although we covaried for age, one possible explanation for the negative findings on clinical symptoms and cognition is that the anodal tDCS group were significantly younger with larger baseline clinical and cognitive impairments compared to sham, both of which could have impaired learning [89]. This would be supported by evidence that ADHD children with worse neurocognitive skills at baseline show less CT gains [90] and neurofeedback learning [91–93], while in healthy controls poorer cognitive performance at baseline can lead to null and even detrimental effects of tDCS [94,95].

Alternatively, given the stronger electric field strengths in children than in adults [96,97], multiple sessions of tDCS may have triggered a homeostatic plasticity response – i.e., the amount and direction of plasticity was attenuated in response to excessive increases in neuronal excitability [98–100] – thereby temporarily disrupting the excitability of rIFC [99–101]. This is in line with our post-hoc moderation analysis that revealed older but not younger participants improved less in the ADHD-RS Total Scores in the anodal versus sham tDCS group at posttreatment. Future studies should verify if tDCS has differential effects depending on current strength and age of participants with ADHD.

Another possibility is that rIFC stimulation downregulated neighbouring dorsal prefrontal or parietal regions part of the dorsal attention network [7,102], or left hemispheric prefrontal regions that mediate positive emotions [103,104].

Interestingly, however, impulsiveness/hyperactivity symptoms, which are most closely associated with rIFC activation [3,105], were lower at follow-up in the anodal relative to the sham tDCS group. The finding – that needs replication – could suggest longer-term neuroplastic consolidation effects as have been shown in other neurotherapies, such as neurofeedback [106,107].

Both groups improved in symptoms and cognitive performance from baseline to posttreatment or follow-up, which could suggest gains due to CT [62]; however, given the lack of correlation with CT performance, placebo effects cannot be ruled out.

The negative findings from this trial are crucial given that tDCS is being increasingly incorporated into clinical practice, is considered an acceptable alternative to medication by parents, and is already commercially available [17,108]. Particularly alarming is that parent-rated ADHD symptoms and adverse effects were higher at posttreatment after anodal tDCS relative to sham.

Findings are not encouraging for the efficacy of multi-session tDCS of rIFC combined with CT in ADHD. However, there are limitations to the study. Although our sample of 50 participants is the largest sample of any tDCS study in children and adolescents with ADHD, larger studies may be more adequately powered to detect effects. We cannot rule out that positive results are achievable with other study designs and stimulation parameters. Computational current flow models suggest that higher stimulation intensities might be required to modulate clinical symptoms and cognitive functions mediated by rIFC given that this it is a deeper region compared to the commonly stimulated dlPFC [97]. Further, this study stimulated “F8” in line with other studies [51–53,109–111]; however, improved performance on inhibitory control tasks in healthy adults has been reported when stimulating T4-Fz and F8-Cz intersection [112–116] or F6 [117–120], which cover the rIFC along with areas closer to the surface implicated in motor inhibition (e.g., superior and middle frontal cortex, and the supplementary motor area) [7,42,121] and attention (e.g., right dlPFC, part of the dorsal attention network and typically underactivated in ADHD) [7,102]. Another limitation is that we could not test for weekly dose effects as ADHD symptoms were only measured at baseline, posttreatment and follow-up; yet weekly changes in ACTIVATE™ game performance and stop task PI did not show dose effects.

Larger, double-blind, randomised-controlled trials should systematically investigate optimal and ideally individualised stimulation protocols (e.g., different stimulation sites, intensity, duration, number of sessions, etc.) measuring clinical, cognitive, and possible non-targeted cognitive outcomes. Stimulating T4-Fz and F8-Cz intersection and F6 could potentially be more effective for improving inhibitory control and attention functions in ADHD.

## Conclusion

This rigorously conducted double-blind, randomised, sham-controlled trial of 15-weekday sessions of anodal tDCS over rIFC combined with CT in 50 boys with ADHD showed no clinical or cognitive improvement. Findings suggest that rIFC stimulation may not be indicated as a neurotherapy for cognitive or clinical remediation for ADHD.

## Data Availability

Anonymized data are available on request

## Abbreviations

ADHD: Attention-Deficit/Hyperactivity Disorder
CT: Cognitive Training
EF: Executive Functions
MARS: Maudsley Attention and Response Suppression
WCST: Wisconsin Card Sorting Task

## CREDIT ROLES

**Conceptualization:** Samuel J. Westwood, Marion Criaud, Sheut-Ling Lam, Steve Lukito, Bruce Wexler, Roi Cohen-Kadosh, Philip Asherson and Katya Rubia. **Data Curation**: Samuel J. Westwood, Sophie Wallace-Hanlon and Afroditi Kostara. **Formal Analysis**: Samuel J. Westwood, Marion Criaud, Sheut-Ling Lam, Steve Lukito, Deborah Agbedjro, Roi Cohen-Kadosh and Katya Rubia. **Funding Acquisition**: Bruce Wexler, Roi Cohen-Kadosh, Philip Asherson and Katya Rubia. **Investigation**: Samuel J. Westwood, Marion Criaud, Sheut-Ling Lam, Steve Lukito, Sophie Wallace-Hanlon, Olivia S. Kowalczyk, Afroditi Kostara and Joseph Mathew. **Methodology**: Samuel J. Westwood, Marion Criaud, Sheut-Ling Lam, Steve Lukito, Olivia S. Kowalczyk, Bruce Wexler, Roi Cohen-Kadosh, Philip Asherson and Katya Rubia. **Project Administration**: Samuel J. Westwood and Katya Rubia. **Resources**: Bruce Wexler, Roi Cohen-Kadosh, Philip Asherson and Katya Rubia. **Software**: Marion Criaud, Bruce Wexler and Katya Rubia. **Supervision**: Samuel J. Westwood and Katya Rubia. **Validation**: Samuel J. Westwood and Katya Rubia. **Visualization**: Samuel J. Westwood and Katya Rubia. **Writing - Original Draft Preparation**: Samuel J. Westwood and Katya Rubia. **Writing - Review & Editing**: Samuel J. Westwood, Marion Criaud, Sheut-Ling Lam, Steve Lukito, Sophie Wallace-Hanlon, Olivia S. Kowalczyk, Deborah Agbedjro, Bruce Wexler, Roi Cohen-Kadosh and Katya Rubia.

## ACKNOWLEDGEMENTS

We would like to thank the families and children who took part in this study, and the South London and Maudsley NHS Trust and local parent groups for their support and participation in this study. We would like to thank Yuanyuan Yang for her help in recruiting participants and data collection.

## Footnotes

1 The saline solution was held in *tufts* by capillary forces between strands, which prevented it from spreading/dripping.

