## Supplement for "Transcranial direct current stimulation (tDCS) combined with cognitive training in adolescent boys with ADHD: a double-blind, randomised, sham-controlled trial"

### **MARS TRAINING STOP TASK**

This task is a visual and faster modification (Rubia et al., 2007) of the tracking stop signal task (Logan et al., 1997). The instructions were purposely changed relative to other versions to make this a training task. The task contains an algorithm which dynamically adjusts in response to whether participants successfully inhibit responses to stop signals (Rubia et al., 2003). In 73% of trials, a spaceship (Go stimulus) pointing left or right is displayed in the centre of the screen for 1000msecs followed by a blank screen for 700msecs, and participants must press the left or right arrow key according to the direction the spaceship is pointing. In 27% of trials, the spaceship is followed by a red explosion presented for 300msecs (stop signal), in which event participants had to inhibit their response to the spaceship. With the intention of training their ability to withhold prepotent responses to Go stimuli (inhibition) and to delay their responses (timing), participants were told that they may have to wait to see if a stop signal appeared after the go stimulus. An algorithm dynamically adjusts the delay between a Go stimulus and a stop signal (i.e., stop signal delay) in steps of 50msecs if successful inhibition moved above or below 50%. The stop signal delay starts at 250msecs. If successful inhibition rose above 50%, the stop signal delay increased by 50ms, making it harder to inhibit a response. If successful inhibition fell below 50%, the stop signal delay reduced by 50msecs, making it easier to inhibit a response. There were 178 trials in total, with 48 stop trials (24 after a left-handed go response and 24 after a right-handed go response). Given that the task was used for training inhibitory control, the participants were asked to try to inhibit as many stop trials as possible and to explicitly wait for the Stop signal to appear to maximise the ability to inhibit their responses. The dependent variable in this training Stop task version is the probability of inhibition.

### **STANDARD OPERATING PROCEDURE FOR COGNITIVE TASKS**

**Task Order**

Participants with ADHD were administered the cognitive tasks in the following order.

1. MARS Go/No-Go task (5 mins )
2. MARS Simon task (5 mins 8 secs)
3. MARS Continuous Performance Task (CPT) (8 mins)
4. MARS Time Discrimination task (4 mins 30 secs)
5. EEG (rest and one Go/No-Go block) (9.5 mins)
6. Mackworth Clock task (MCT) (6 mins)
7. Wisconsin Card Sorting task (WCST) (20 mins)
8. NIH Toolbox List Sorting Working Memory (WM) task (at least 5 mins)
9. Verbal Fluency task (7 mins)

For tasks 1 to 8, the experimenter gave instructions with visual aids and performed the task as a demonstration. Participants then practiced the task until the experimenter was satisfied that the instructions were understood. All tasks were performed on a Dell XPS 15.6 Inch QHD Touch Laptop.

**Task Protocols**

**Maudsley Attention and Response Suppression (MARS) tasks.** All tasks required participants to press the left or right arrow key on the keyboard with their corresponding index finger in accordance with task instructions (Rubia et al., 2007; Schmitz et al., 2008; Penadés et al., 2007).

*MARS Go/No-Go (GNG).* A measure of motor response inhibition, GNG requires a motor response to Go stimuli and response inhibition to No-Go stimuli. The task was split into two 2.30 minute blocks, blocked for a left- or a right-handed response only in each block to increase prepotent response tendency. Participants always started with the left-handed response.

- **Block 1, left-handed response**: In 73.4% of trials, a spaceship (Go stimulus) pointing left appears in the centre of the screen and participants must press the left arrow key as fast as possible. In 26.6% of trials, a blue planet (No-Go stimulus) appears in the centre of the screen instead of a spaceship and participants must inhibit their response.
- **Block 2, right-handed response**: Block 2 was identical to Block 1, except spaceships face right and participants respond using the right arrow key.

Go and No-Go stimuli are displayed for 300msecs followed by a blank screen for 1000msecs. There are 300 trials in total (2 x 110 Go trials, 2 x 40 No-Go trials). The dependent variable is the probability of inhibition to No-Go stimuli.

*MARS Continuous Performance Task (CPT)***.** This target detection task measures sustained attention. Individual letters (A to L, O and X) are displayed consecutively and participants have to press the left arrow key when “A” was followed by an “X” or the right arrow key when “A” was followed immediately by an “O”. Participants do not respond to any of the other letters or letter combinations. Feedback is given via a progress bar composed of red or blue coloured boxes numbered 1 to 10, which are presented at the right-hand side of the screen. For every three correct responses, a new box lights up, blue for A-O trials or red for A-X trials. Participants were told that the goal was to get all of the boxes in both bars to light up, indicating that all 60 trials had been successfully responded to.

Letters were displayed for 300msecs followed by a blank screen of 700msecs. There were 480 trials in total, with 60 target letter trials (12.5%) of which half were “A” followed by “X” and the other half were “A” followed by “O”. The dependent variables were the percentage of omission errors on target trials and percentage of commission errors (i.e. false hits) on nontarget trials.

*Simon task***.** This task measured stimulus-response conflict resolution/interference inhibition and selective attention. In this task, arrows pointing left or right appear on the left- or right-hand side of the screen. Participants must press the arrow key that corresponds to the direction the arrow is pointing as fast as they can. In 72.73% of trials, the direction an arrow pointed and the side of the screen it appeared is *congruent* (e.g., left arrows appears on the left, and vice versa); the remaining 27.27% trials are *incongruent* trials (e.g., left arrows appeared on the right, or right arrows on the left). Response conflict arises between iconic information (i.e., a left-hand response to a left-pointing arrow) and the predominant, incompatible spatial information (i.e., the arrow appears on the opposite side of the screen it is pointing toward, e.g., right). This conflict is typically reflected in slower reaction times to incongruent relative to congruent trials and the difference between these trials (RT incongruent – RT congruent) is called the Simon reaction time effect (Simon & Berbaum, 1988)

Arrows were displayed and then followed by a blank screen with an inter stimulus interval of 1400msecs. There were 220 trials in total, 160 congruent trials (80 left arrows, 80 right arrows) and 60 incongruent trials. The dependent variable is the Simon RT effect (i.e., RT incongruent – RT congruent, the Simon RT effect).

*Time discrimination task*. In this time discrimination task, participants have to discriminate between time intervals of 1s durations that differ by several hundred of milliseconds. A red and a green circle (5 cm in diameter) appears consecutively with no interspersed pause. The red circle appears on the left side and the green circle on the right side of the screen.

One circle is randomly presented for a standard duration of 1 s, and the other circle for either 1.3 s, 1.4 s, or 1.5secs (the total trial times were either 4.4, 4.5, or 4.6 s) followed by a blank screen presented for 2.1 s in which time participants make their response. Participants had to decide which of the two circles appeared on the screen for the longest time, pressing either the right arrow for the red circle or the left arrow for the green. Participants were instructed to count the time in order to make their decision. Sixty pairs of stimuli were presented. The dependent variable was the percentage of total correct response.

*Mackworth Clock* (Lichstein et al., 2000; Mackworth, 1948; PsyTookit, 2017). This vigilance task measures identification of difficult to detect signals. A clockface was displayed on the screen with a secondhand that ‘ticked’ around the face every second. At infrequent and irregular intervals, the secondhand ‘skipped’ a second, at which point participants had to press the space bar as soon as possible. The hand skipped in 10% of trials (40 out of 400 ticks around the face) in a pseudorandomised order that was the same across participants. The dependent variables were the percentage of omission and commission errors.

*Wisconsin Card Sorting Task* (WCST, PsyToolkit, 2018)*.* The WCST measures cognitive flexibility/switching. Participants are shown a deck of 64 cards and a row of four cards on the computer screen. They were told to match each card from the deck with one of the four cards and that the cards could be matched by colour, shape or number of symbols depicted on the cards. Participants responded by tapping on one of the four cards at the top of the screen using the laptop touch screen. Verbal and visual feedback informed participants if their match was correct or incorrect. After 10 consecutive trials, the sorting rule changed without the participant being informed and the participant had to switch to a new rule.

There were 60 trials (i.e., 6 rule changes) in total. The dependent variables were the total number of perseverative errors (i.e., where the rule changed to a new one but the participant continued to match cards using the old rule) and non-perseverative errors (i.e., where the rule changed to a new one and the participant match cards using the new rule).

*NIH List Sorting Test Working Memory task* (see Tulsky et al., 2014)*.* In this measure of visuo-spatial working memory, a sequence of pictures is displayed followed by a 3 X 4 matrix of the same and new pictures. Participants select the pictures presented in the sequence in size order from smallest to largest. The task is split into two blocks, grouped by stimuli type.

- **Block 1.** Participants were presented sequences of pictures of either animals or objects. The task started with a sequence of two pictures, which increased by one picture (up to seven) after every correct trial. If a response was incorrect, the same number of pictures was presented. The block was automatically terminated in the event of incorrect responses on two trials with the same number of pictures or a correct response to one trial with seven pictures. Block 1 was followed immediately by Block 2.
- **Block 2.** Block 2 was identical to Block 1 except both animals *and* objects were presented in the same sequence.

The dependent variable was total number of correct responses across both blocks.

*Verbal Fluency.* This task is a standard measure of language production (Troyer, 2000). Participants were given 60 seconds to name as many unique words as possible that started with a given letter (C, L, F) or belonged to a give semantic category (Fruits, Super Market Items, and Animals). Proper names (e.g., Rochester or Robert) or repetitions (even with a different ending; e.g., eat followed by eating) were not allowed. Participants were reminded to keep going until the time ran out even if they drew a blank. To ensure participants understood the task, the experimenter provided a practice example (e.g., for the letter “T”, I could say, “terrible,” “turn”, and “table”) and asked participants if they could think of any other words.

Responses were scored after the testing session. Slang words and foreign words were permissible answers so long as they were listed as standard English words. Participants were asked to indicate the meaning of a word in instances of ambiguity (e.g., frank) at the end of the task. The dependent variable is the percentage of unique, correct words.

### **GENERIC EXECUTIVE FUNCTIONING MEASURES**

We tested the effect of tDCS and cognitive training on three generic, task-independent executive function (EF) measures, which included general speed of information processing (mean reaction times [MRT], response variability (coefficient of variance [CV]), and premature/impulsive responses across Go/No-Go, Simon, CPT Tasks.

**Testing for correlations between measures.**

Before combining these generic executive function measures, we correlated MRT, CV, and premature responses at baseline across GNG, Simon, and CPT. Our findings indicated fair to good (i.e., Pearson’s *r* and Spearman’s *rho* > 0.3) statistically significant correlations at baseline for MRTs, CV, and premature responses (see Tables 1 to 3 and Figures 1 to 3).

**Outcome Calculation**

**Speed of information processing.** We combined MRT for GNG Go Trials, Simon Congruent Trials, and CPT Target Trials. Because the number of trials differed for each task, MRT were weighted by multiplying MRT by the weighting factor (i.e., the total number of trials across tasks divided by the number of trials in a given task) (see Table 4). Weighted MRT were summed to create the combined MRT.

**Intrasubject response variability**. We combined the Intrasubject CV for GNG Go Trials, Simon Congruent Trials, and CPT Target Trials. CV provides a measure of RT variability while controlling for an individuals’ overall speed of RT (Rubia et al., 2007). For each participant, CV was calculated using the following formula SD of MRT/MRT - i.e.,   standard deviation of the MRT divided by the MRT.

**Premature responses***.* We combined premature responses to all GNG, Simon, and CPT trials. For all tasks, we defined premature responses as responses made 200ms before and 100ms after stimulus onset. This is because 200ms before stimulus onset is too early for the type of stimulus to be seen by the participant, while 100ms after stimulus onset is too short for a normal reaction time (Rubia et al., 2007). Because the number of trials differed for each task, premature responses were weighted by multiplying premature response by the weighting factor (Table 5). Weighted premature responses were summed to create the combined measure of prematurity.

### **SENSITIVITY ANALYSES**

We repeated our confirmatory analysis of primary and secondary outcome measures excluding a) statistical outliers in cognitive measures, b) participants who changed medication between posttreatment and follow-up, or c) participants whose tDCS and CT sessions extended over four weeks.

**Statistical Outliers**

Studentized Residuals were calculated from repeated measures ANCOVA testing group differences in primary and secondary outcomes at posttreatment and at follow-up separately, adjusting for baseline, age in months, and medication status (Naïve, On- or Off-medication). We defined an outlier as a Studentized Residual that was 3 times greater than the interquartile range (IQR) when combining data from both anodal and sham tDCS groups. This resulted in excluding several participants from several cognitive tasks (see Table 5).

**Medication Status Change**

Our protocol requested that participants remain on stable medication from baseline to posttreatment, but not to follow-up as there are ethical concerns in keeping participants on a fixed medication regime longer than is necessary. Two participants from the anodal tDCS group changed their medication status from posttreatment and follow-up (one ceased taking medication entirely, the other went from being medication-naïve to taking stimulant medication). Given this change in medication status could confound our measures of the clinical and cognitive effects of tDCS, we ran a sensitivity analysis excluding these individuals (see Table 6 and 7).

**Extra Consolidation**

Any session that could not be attended was rescheduled for the first weekday available after the remaining sessions had been completed. This occurred for six participants, with five rescheduled for the first available Monday and one for the first available Tuesday. Because tDCS effects can be potentiated with a weekend interval due to extra consolidation time (Au et al., 2016, 2017), participants whose sessions were rescheduled had an extra weekend of consolidation and therefore could potentially show larger effects relative to other participants. To assess the influence of these individuals on our results, we ran a sensitivity analysis excluding participants who had an extra weekend of consolidation (see Table 6 and 7).

| Table 1. Pearson’s r and Spearman’s rho correlations between MRTs in GNG Go-Trials, Simon Congruent Trials, and CPT Target trials at baseline | | | | |
| --- | --- | --- | --- | --- |
|  |  | GNG Go Trials | Simon Congruent Trials | CPT Target Trials |
| GNG Go Trials | Pearson's r | — |  |  |
|  | p-value | — |  |  |
|  | Spearman's rho | — |  |  |
|  | p-value | — |  |  |
| Simon Congruent Trials | Pearson's r | 0.540 | — |  |
|  | p-value | < .001 | — |  |
|  | Spearman's rho | 0.532 | — |  |
|  | p-value | < .001 | — |  |
| CPT Target Trials | Pearson's r | 0.270 | 0.476 | — |
|  | p-value | 0.058 | < .001 | — |
|  | Spearman's rho | 0.374 | 0.460 | — |
|  | p-value | 0.008 | < .001 | — |

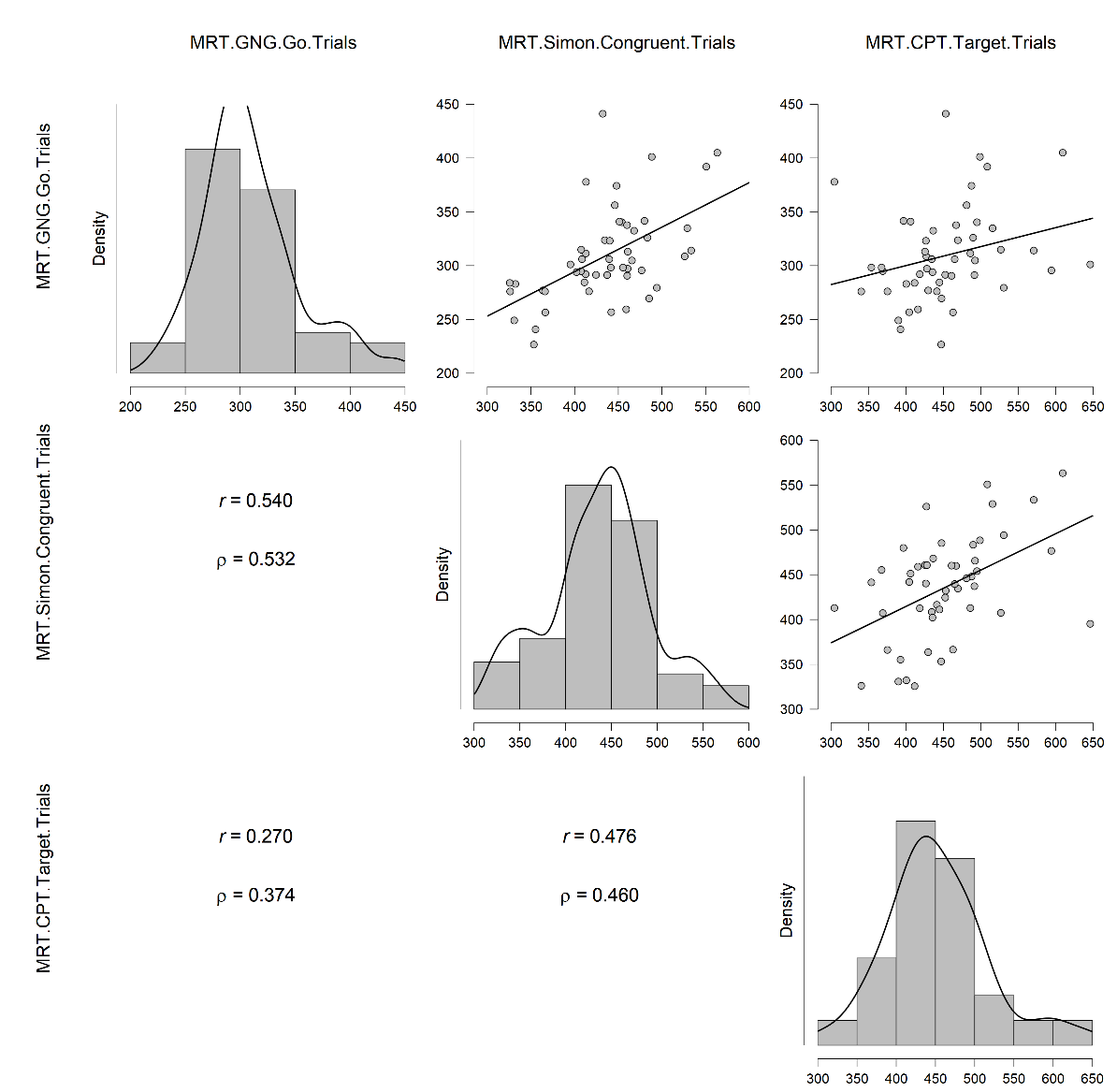

[Figure](https://docs.google.com/document/d/1w9EUaPH0tm6FIAH1q1a7W55bKQUCzk4A4POmM-4he9k/edit#figur_gng_comb) 1. Correlations between MRT in GNG Go Trials, Simon Congruent Trials, and CPT Target Trials.

| Table 2. Correlations between CV in GNG Go-Trials, Simon Congruent Trials, and CPT Target Trials | | | | |
| --- | --- | --- | --- | --- |
|  |  | CV Go Trials | CV Simon Congruent Trials | CV CPT Target Trials |
| CV GNG Go Trials | Pearson's r | — |  |  |
|  | p-value | — |  |  |
|  | Spearman's rho | — |  |  |
|  | p-value | — |  |  |
| CV Simon Congruent Trials | Pearson's r | 0.767 | — |  |
|  | p-value | < .001 | — |  |
|  | Spearman's rho | 0.733 | — |  |
|  | p-value | < .001 | — |  |
| CV CPT Target Trials | Pearson's r | 0.538 | 0.592 | — |
|  | p-value | < .001 | < .001 | — |
|  | Spearman's rho | 0.460 | 0.518 | — |
|  | p-value | < .001 | < .001 | — |

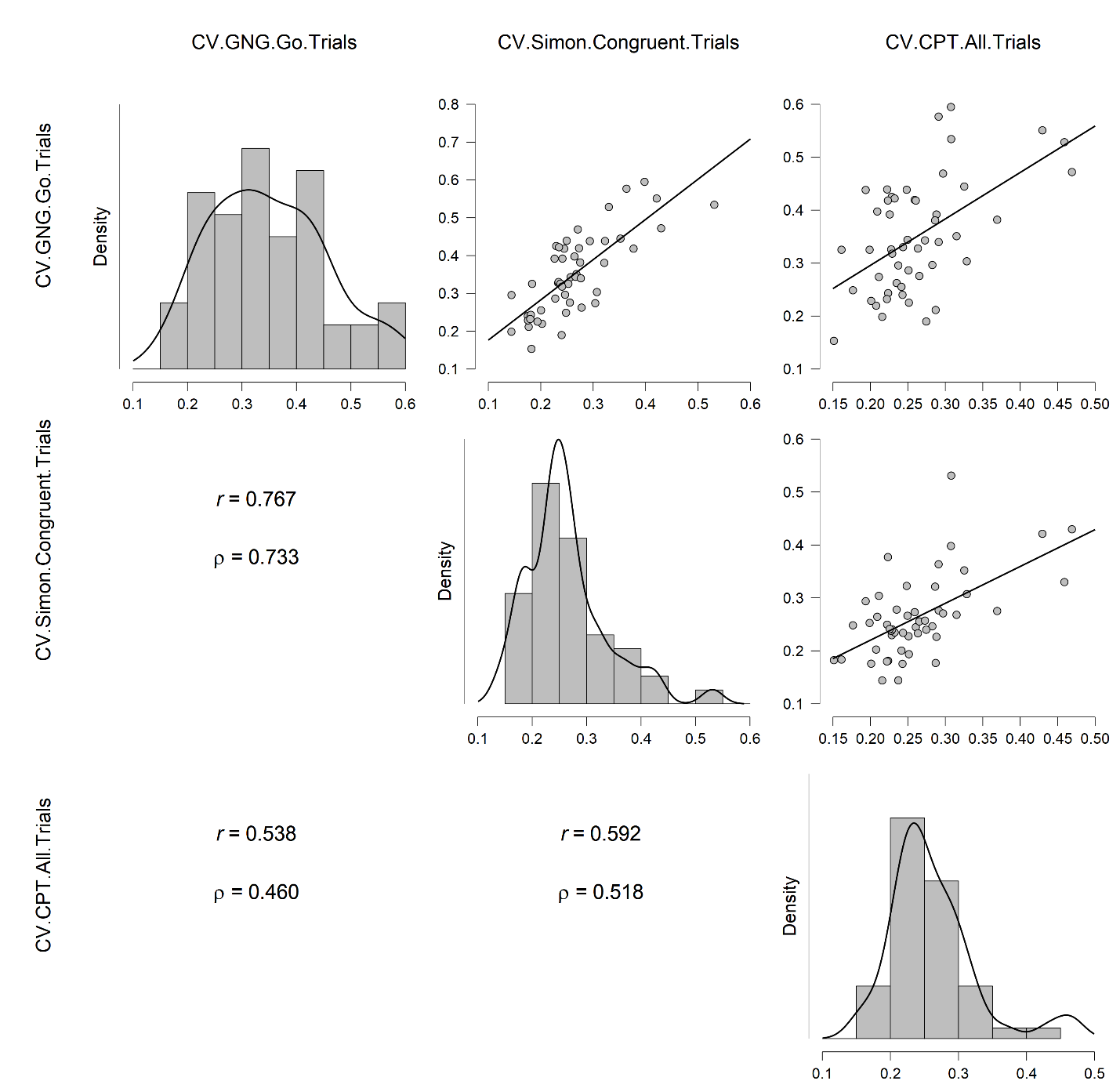

[Figure](https://docs.google.com/document/d/1w9EUaPH0tm6FIAH1q1a7W55bKQUCzk4A4POmM-4he9k/edit#figur_gng_comb) 2. Correlations between CV in GNG Go Trials, Simon Congruent Trials, CPT Target Trials

| [Table](https://docs.google.com/document/d/1w9EUaPH0tm6FIAH1q1a7W55bKQUCzk4A4POmM-4he9k/edit#table_prem) 3. Correlations between percentage of premature responses in all GNG, Simon and CPT Trials ls) | | | | |
| --- | --- | --- | --- | --- |
|  |  | Premature % GNG Trials | Premature % Simon Trials | Premature % CPT Trials |
| Premature % GNG Trials | Pearson's r | — |  |  |
|  | p-value | — |  |  |
|  | Spearman's rho | — |  |  |
|  | p-value | — |  |  |
| Premature % Simon Trials | Pearson's r | 0.531 | — |  |
|  | p-value | < .001 | — |  |
|  | Spearman's rho | 0.624 | — |  |
|  | p-value | < .001 | — |  |
| Premature % CPT Trials | Pearson's r | 0.601 | 0.676 | — |
|  | p-value | < .001 | < 0.001 | — |
|  | Spearman's rho | 0.452 | 0.671 | — |
|  | p-value | < .001 | < .001 | — |

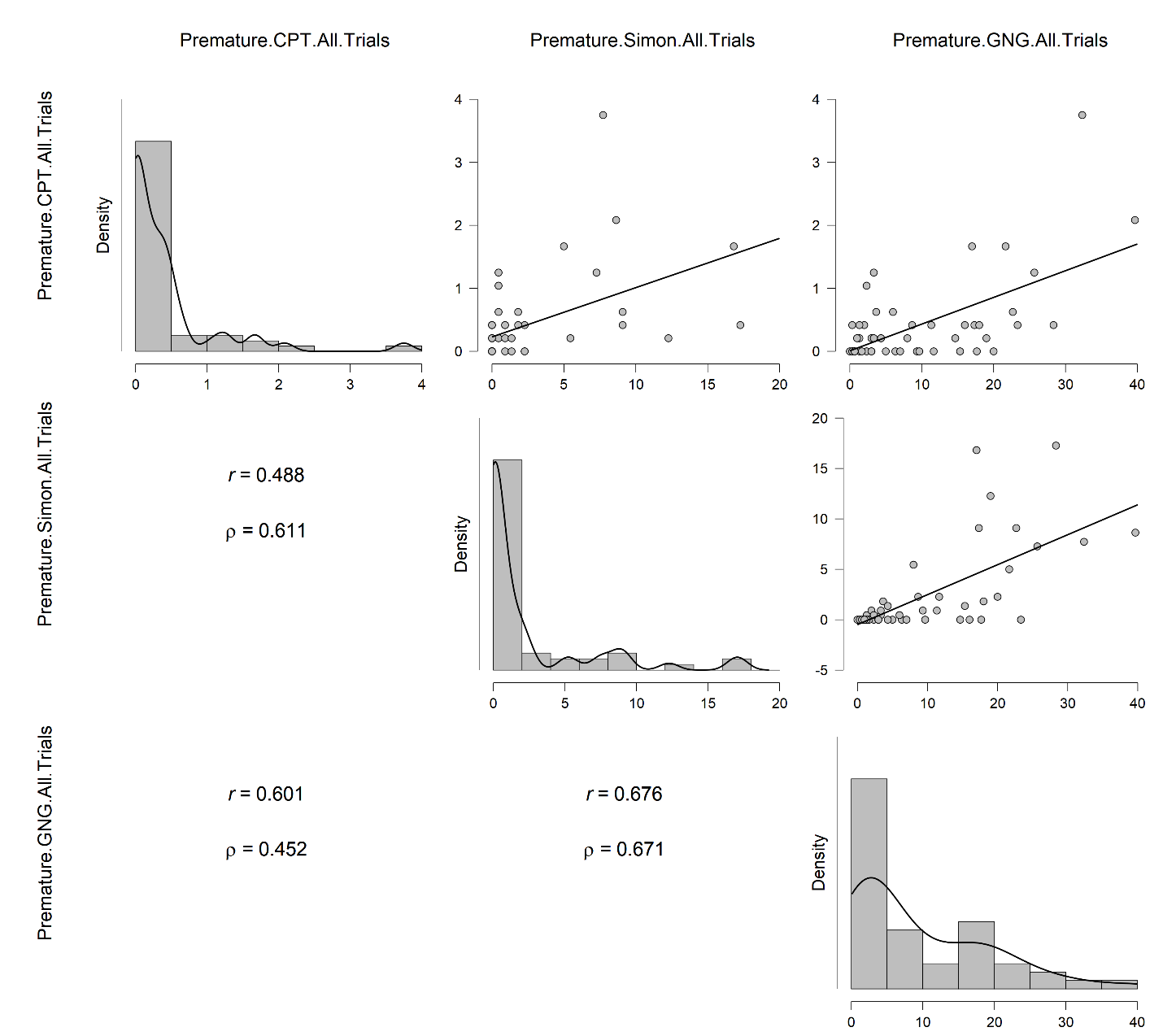

[Figure 3](https://docs.google.com/document/d/1w9EUaPH0tm6FIAH1q1a7W55bKQUCzk4A4POmM-4he9k/edit#figur_prem). Correlations between % of premature responses in all GNG, Simon, CPT Trials

| Table 4. Weighting factor according to the number of trials for each task | | |
| --- | --- | --- |
| **Trial Type** | **Trial No.** | **Weighting Factor (w)*** |
| GNG Go | 220 | 0.50 |
| Simon Congruent | 160 | 0.36 |
| CPT Target | 60 | 0.14 |
| **Sum Total** | 440 | 1.00 |
| *, 440 divided by no. trials in a given task | | |

| Table 5. Weighting factor according to the number of trials for each task | | |
| --- | --- | --- |
| **All Trials** | **Trial No.** | **Weighting Factor (w)*** |
| GNG | 220 | 0.24 |
| Simon | 220 | 0.24 |
| CPT | 480 | 0.52 |
| **Sum Total** | 920 | 1.00 |
| *920 divided by no. trials in a given task | | |

| Table 6. Sensitivity analysis testing group differences on cognitive outcome measures without statistical outliers. | | | | | | | |
| --- | --- | --- | --- | --- | --- | --- | --- |
|  | |  | | **ANCOVA^*^** | |  | |
|  | | **Time** | | **Group** | | **Group by Time** | |
|  | ***N*** | ***F (1,42)*** | ***p†*** | ***F (1,42)*** | ***p†*** | ***F (1,42)*** | ***p†*** |
| ***CPT: Omission (%)*** |  |  |  |  |  |  |  |
| *Anodal tDCS* | 23 | .36 | .55 (1.00) | 5.70 | **.02 (.04)** | .09 | .77 (.77) |
| *Sham tDCS* | 25 |  |  |  |  |  |  |
| ***CPT: Commission (%)*** |  |  |  |  |  |  |  |
| *Anodal tDCS* | 23 | .004 | .95 (.95) | 4.13 | **.05** (**.05**) | .36 | .55 (1.00) |
| *Sham tDCS* | 25 |  |  |  |  |  |  |
| ***Fluency: Letter % Corr*** |  |  |  |  |  |  |  |
| *Anodal tDCS* | 23 | .37 | .55 (.92) | 5.37 | **.03** (.15) | 2.58 | .12 (.30) |
| *Sham tDCS* | 25 |  |  |  |  |  |  |
| ***Fluency: Semantic % Corr*** |  |  |  |  |  |  |  |
| *Anodal tDCS* | 23 | .13 | .72 (.90) | .52 | .48 (.48) | 3.15 | .08 (.40) |
| *Sham tDCS* | 25 |  |  |  |  |  |  |
|  |  | ***F (1,43)*** | ***p*** | ***F (1,43)*** | p | ***F (1,43)*** | p |
| ***Macworth Task: Commissions (%)*** |  |  |  |  |  |  |  |
| *Anodal tDCS* | 24 | 3.87 | **.06** (.30) | 1.92 | .17 (.21) | .06 | .81 (.81) |
| *Sham tDCS* | 25 |  |  |  |  |  |  |
| ***Macworth Task: Omissions (%)*** |  |  |  |  |  |  |  |
| *Anodal tDCS* | 24 | 1.43 | .24 (.60) | 3.10 | .09 (.15) | .144 | .71 (.89) |
| *Sham tDCS* | 25 |  |  |  |  |  |  |
| ***Prematurity: Premature Responses*** |  |  |  |  |  |  |  |
| *Anodal tDCS* | 24 |  |  |  |  |  |  |
| *Sham tDCS* | 25 | .014 | .91 (.91) | 3.66 | .06 (.15) | 1.83 | .18 (.30) |
| *Repeated measures ANCOVA testing group differences at posttreatment and follow-up (where applicable), adjusting for baseline, age at entry and medication status (Naïve, Off-medication, On-medication)  ***†***Benjamini-Hochberg adjustment was applied to p-values for time, group, and time by group interaction effect separately and was applied separately to primary cognitive, secondary cognitive, and secondary clinical outcome measures separately. | | | | | | | |

| Table 7. Sensitivity analysis testing group differences on primary outcomes excluding two participants who changed their medication status between posttreatment to follow-up or participants who had an extra weekend of consolidation. Benjamini-Hochberg adjusted p-values in parentheses. | | | | | | | |
| --- | --- | --- | --- | --- | --- | --- | --- |
|  |  | **ANCOVA*** | | | | | |
|  |  | **Time** | | **Group** | | **Group by Time** | |
| ***Sensitivity analysis*** | ***Measure*** | ***F (1,42)*** | ***p†*** | ***F (1,42)*** | ***p†*** | ***F (1,42)*** | ***p†*** |
| *Excluding participants with*  *a change in medication status* | ***Go/No-Go*** |  |  |  |  |  |  |
|  | *PI (%)* | .30 | .59 (1.00) | .22 | .65 (.65) | .07 | .79 (.79) |
|  | ***CPT*** |  |  |  |  |  |  |
|  | *Omission (%)* | .19 | .67 (1.00) | 3.93 | **.05 (.15)** | .16 | .69 (1.00) |
|  | *Commission (%)* | .01 | .94 (.94) | 3.23 | .08 (.12) | .84 | .36 (1.00) |
|  | ***ADHD RS*** |  |  |  |  |  |  |
|  | *Total Score* | 1.37 | .25 | .18 | .67 | 9.40 | **.004** |
|  |  | ***F (1,38)*** | ***p*** | ***F (1,38)*** | ***p*** | ***F (1,38)*** | ***p*** |
| *Excluding participants with*  *an extra weekend of consolidation* | ***Go/No-Go*** |  |  |  |  |  |  |
|  | *PI (%)* | .001 | .98 (.98) | .03 | .87 (.87) | .03 | .87 (.87) |
|  | ***CPT*** |  |  |  |  |  |  |
|  | *Omission (%)* | .95 | .34 (1.00) | 4.06 | **.05** (.08) | .41 | .52 (1.00) |
|  | *Commission (%)* | .05 | .83 (1.00) | 4.32 | **.04** (.12) | .28 | .60 (.90) |
|  | ***ADHD RS*** |  |  |  |  |  |  |
|  | *Total Score* | 1.21 | .28 | .80 | .38 | 3.37 | **.03** |
| *Repeated measures ANCOVA testing group differences at posttreatment and follow-up (where applicable), adjusting for baseline, age at entry and medication status (Naïve, Off-medication, On-medication)  ***†***Benjamini-Hochberg adjustment was applied to p-values for time, group, or time by group interaction effects separately and was applied separately to primary cognitive, secondary cognitive, and secondary clinical outcome measures separately. | | | | | | | |

| Table 8. Sensitivity analysis testing group differences on secondary outcome measures excluding two participants who changed their medication status between posttreatment to follow-up. Benjamini-Hochberg adjusted p-values in parentheses. | | | | | | | | | | | | | | | | |
| --- | --- | --- | --- | --- | --- | --- | --- | --- | --- | --- | --- | --- | --- | --- | --- | --- |
|  | | | | |  | | | | | **ANCOVA*** | | | |  | | |
|  | | | | |  | | | | | **Group** | | | | **Group by Time** | | |
| ***Sensitivity analysis*** | | | ***Measure*** | | ***F (1,42)*** | | ***p†*** | | | ***F (1,42)*** | | | ***p†*** | ***F (1,42)*** | | ***p†*** |
| *Excluding participants with*  *a change in medication status* | | | ***Simon*** | |  | |  | |  | | | |  |  | |  |
|  |  |  | Simon RT Effect | | .01 | | .92 (1.00) | | | 3.16 | | | .08 (.96) | .23 | | .63 (1.00) |
|  |  |  | ***Time Discrimination*** | |  | |  | | |  | | |  |  | |  |
|  | | | Total Correct (%) | | 3.51 | | .07 (.84) | | | 1.13 | | | .29 (.58) | .11 | | .74 (.99) |
|  | | | ***Macworth Clock*** | |  | |  | | |  | | |  |  | |  |
|  | | | Commissions (%) | | 2.07 | | .16 (.96) | | | .04 | | | .85 (.85) | .12 | | .73 (1.00) |
|  | | | Omissions (%) | | 1.41 | | .24 (.72) | | | 1.75 | | | .19 (.76) | .81 | | .37 (1.00) |
|  | | | ***WCST*** | |  | |  | | |  | | |  |  | |  |
|  | | | Non Perseveration | | 1.81 | | .19 (.76) | | | .77 | | | .39 (.67) | .52 | | .47 (1.00) |
|  | | | Pers Errors | | .14 | | .71 (.95) | | | .08 | | | .77 (.84) | .41 | | .53 (1.00) |
|  | | | ***List sort WM*** | |  | |  | | |  | | |  |  | |  |
|  | | | Total Score | | .03 | | .86 (1.00) | | | .23 | | | .63 (.84) | .10 | | .75 (.90) |
|  | | | ***Fluency*** | |  | |  | | |  | | |  |  | |  |
|  | | | Letter % Corr | | .44 | | .51 (.87) | | | 1.43 | | | .24 (.58) | 1.71 | | .20 (1.00) |
|  | | | Semantic % Corr | | .15 | | .70 (1.00) | | | 1.50 | | | .23 (.69) | 5.43 | | .03 (.36) |
|  | | | ***Speed of Processing*** | |  | |  | | |  | | |  |  | |  |
|  | | | MRT | | 1.28 | | .27 (.54) | | | .59 | | | .45 (.68) | .01 | | .95 (.95) |
|  | | | ***Response Variability*** | |  | |  | | |  | | |  |  | |  |
|  | | | CV | | 1.39 | | .25 (.60) | | | .11 | | | .74 (.89) | .03 | | .86 (.94) |
|  | | | ***Prematurity*** | |  | |  | | |  | | |  |  | |  |
|  | | | Premature Resp. | | <.001 | | .99 (.99) | | | 2.24 | | | .14 (.84) | 1.28 | | .26 (1.00) |
|  | | | ***Conners 3-P ADHD Index*** | | .004 | | .95 (.95) | | | 1.74 | | | .19 (.57) | 13.80 | | .001 (.02) |
|  | | | ***ARI*** | |  | |  | | |  | | |  |  | |  |
|  | | | *Parent* | | .49 | | .98 (.49) | | | 1.42 | | | .24 (.51) | 3.13 | | .08 (.16) |
|  | | | *Child* | | .21 | | .87 (.65) | | | .09 | | | .77 (.77) | .05 | | .82 (.82) |
|  | | | ***MEWS*** | | 3.32 | | .32 (.08) | | | .62 | | | .44 (.53) | 1.39 | | .24 (.32) |
|  | | | ***WREMB-R*** | | n/a | | n/a | | | 2.14 | | | .15 (.90) | n/a | | n/a |
|  | | | ***CIS*** | | n/a | | n/a | | | .93 | | | .34 (51) | n/a | | n/a |
|  | | |  | | ***F (1,38)*** | |  | | | ***F (1,38)*** | | |  | ***F (1,38)*** | |  |
| *Excluding participants with*  *an extra weekend of consolidation* | | | ***Simon*** | |  | |  | | |  | | |  |  | |  |
|  |  |  | Simon RT Effect | | .06 | | .81 (.81) | | | 3.75 | | | .06 (.24) | .03 | | .86 (1.00) |
|  | | | ***Time Discrimination*** | |  | |  | | |  | | |  |  | |  |
|  | | | Total Correct (%) | | 2.15 | | .15 (1.00) | | | 4.22 | | | **.05** (.30) | .003 | | .96 (.96) |
|  | | | ***Macworth Clock*** | |  | |  | | |  | | |  |  | |  |
|  | | | Commissions (%) | | 1.35 | | .25 (.50) | | | .54 | | | .47 (.63) | .54 | | .47 (1.00) |
|  | | | Omissions (%) | | 1.80 | | .19 (.76) | | | 1.32 | | | .26 (.52) | .44 | | .51 (1.00) |
|  | | | ***WCST*** | |  | |  | | |  | | |  |  | |  |
|  | | | Non Perseveration | | 2.07 | | .16 (.96) | | | .61 | | | .44 (.66) | .71 | | .41 (1.00) |
|  | | | Pers Errors | | .48 | | .50 (.67) | | | >.001 | | | 1.00 (1.00) | .10 | | .75 (1.00) |
|  | | | ***List sort WM*** | |  | |  | | |  | | |  |  | |  |
|  | | | Total Score | | .12 | | .73 (.80) | | | .003 | | | .95 (1.00) | .45 | | .51 (1.00) |
|  | | | ***Fluency*** | |  | |  | | |  | | |  |  | |  |
|  | | | Letter % Corr | | .78 | | .38 (.57) | | | 4.75 | | | **.04** (.48) | 1.07 | | .31 (1.00) |
|  | | | Semantic % Corr | | .91 | | .35 (.60) | | | 2.38 | | | .13 (.31) | 5.6 | | **.02** (.24) |
|  | | | ***Speed of Processing*** | |  | |  | | |  | | |  |  | |  |
|  | | | MRT | | 1.54 | | .22 (.53) | | | .81 | | | .38 (.65) | .14 | | .71 (1.00) |
|  | | | ***Response Variability*** | |  | |  | | |  | | |  |  | |  |
|  | | | CV | | 1.71 | | .20 (.60) | | | .42 | | | .52 (.61) | .02 | | .90 (.98) |
|  | | | ***Prematurity*** | |  | |  | | |  | | |  |  | |  |
|  | | | Premature Resp. | | .26 | | .61 (.73) | | | 2.47 | | | .12 (.36) | .41 | | .52 (.89) |
|  | | | ***Conners 3-P ADHD Index*** | | .08 | | .77 (.77) | | | 3.73 | | | .06 (.36) | 9.66 | | **.004 (.02)** |
|  | | | ***ARI*** | |  | |  | | |  | | |  |  | |  |
|  | | | *Parent* | | .85 | | .36 (.72) | | | 2.72 | | | .11 (.33) | 2.72 | | .11 (.22) |
|  | | | *Child* | | .12 | | .73 (.97) | | | .05 | | | .83 (.83) | .15 | | .70 (.70) |
|  | | | ***MEWS*** | | 3.65 | | .06 (.24) | | | .63 | | | .43 (.52) | 1.43 | | .24 (.32) |
|  | | | ***WREMB-R*** | | n/a | | **n/a** | | | 1.88 | | | .18 (.27) | n/a | | n/a |
|  | | | ***CIS*** | | n/a | | n/a | | | 1.96 | | | .17 (.34) | n/a | | n/a |
| *Repeated measures ANCOVA testing group differences at posttreatment and follow-up (where applicable), adjusting for baseline, age at entry and medication status (Naïve, Off-medication, On-medication)  ***†***Benjamini-Hochberg adjustment was applied to p-values for time, group, and time by group interaction effect separately and was applied separately to primary cognitive, secondary cognitive, and secondary clinical outcome measures separately. | | | | | | | | | | | | | | | | |
| Table 9. Summary of raw average performance on secondary cognitive and clinical outcomes in sham and anodal tDCS groups at baseline, posttreatment, and follow-up. Benjamini-Hochberg adjusted p-values in parentheses. | | | | | | | | | | | | | | | | |
|  |  |  | | ***Unadjusted Mean (SD)*** | | | | | | |  |  | |  |  | |
|  |  |  | | **Baseline** | | **Posttreatment** | | **Follow-up** | | | **Baseline vs**  **Posttreatment** | | | **Baseline vs**  **Follow-Up** | | |
| **Primary Outcomes** | |  | |  | |  | |  | | | ***F(1,48)*** | ***p†*** | | ***F(1,48)*** | ***p†*** | |
| ***Go/No-Go*** | *% PI* | Sham tDCS | | 49.81 (3.82) | | 55.53 (3.82) | | 59.23 (4.28) | | | **9.28** | **.004 (.01)** | | **16.57** | **<.001 (.003)** | |
|  |  | Anodal tDCS | | 46.20 (4.84) | | 54.01 (3.90) | | 55.94 (4.97) | | |  |  | |  |  | |
| ***CPT*** | *% Omission* | Sham tDCS | | 13.59 (2.88) | | 9.62 (1.88) | | 8.33 (1.97) | | | 2.59 | .114 (.14) | | **7.72** | **.01 (.02)** | |
|  |  | Anodal tDCS | | 15.69 (2.81) | | 14.65 (2.26) | | 11.67 (1.75) | | |  |  | |  |  | |
|  | *% Commission* | Sham tDCS | | 1.89 (.62) | | .96 (.20) | | .91 (.22) | | | 2.95 | .09 (.11) | | **8.73** | **.01 (.01)** | |
|  |  | Anodal tDCS | | 2.67 (.56) | | 2.24 (.45) | | 1.33 (.25) | | |  |  | |  |  | |
| ***ADHD RS*** | *Total Score* | Sham tDCS | | 37.08 (1.40) | | 24.31 (1.65) | | 29.42 (1.70) | | | **52.11** | **<.001** | | **51.61** | **<.001** | |
|  |  | Anodal tDCS | | 41.71 (1.67) | | 33.83 (2.04) | | 30.25 (2.74) | | |  |  | |  |  | |
| **Secondary Outcomes** | |  | |  | |  | |  | | |  |  | |  |  | |
| ***Simon*** | Simon RT Effect | Sham tDCS | | 63.4 (30.4) | | 45.3 (24.3) | | 48.6 (20.4) | | | **8.36** | **.006 (.02)** | | **8.06** | **.01 (.02)** | |
|  |  | Anodal tDCS | | 80.2 (45.3) | | 60.4 (36.9) | | 60.3 (29.9) | | |  |  | |  |  | |
| ***Time Discrimination*** | % Total Correct | Sham tDCS | | 76.8 (15.8) | | 73.9 (18.7) | | 80.8 (13.4) | | | 4.15 | .05 (.09) | | 2.92 | .09 (.14) | |
|  |  | Anodal tDCS | | 68.5 (12.8) | | 62.2 (16.7) | | 70.3 (14.6) | | |  |  | |  |  | |
| ***Macworth Clock*** | % Commissions | Sham tDCS | | 3.9 (6.4) | | 3.8 (5.5) | | 4.55 (10.2) | | | 2.74 | .10 (.13) | | .95 | .33 (.33) | |
|  |  | Anodal tDCS | | 8.6 (9.0) | | 5.7 (5.0) | | 5.4 (5.5) | | |  |  | |  |  | |
|  | % Omissions | Sham tDCS | | 34.4 (18.5) | | 29.3 (16.2) | | 23.8 (14.1) | | | 5.97 | .02 (.06) | | **25.04** | **<.001 (.01)** | |
|  |  | Anodal tDCS | | 49.3 (18.0) | | 44.2 (23.2) | | 37.8 (14.4) | | |  |  | |  |  | |
| ***WCST*** | Non-Perseverative | Sham tDCS | | 8.31 (4.19) | | 7.39 (5.14) | | 7.04 (4.02) | | | .001 | .98 (.98) | | 1.19 | .28 (.31) | |
|  |  | Anodal tDCS | | 7.67 (3.74) | | 8.63 (4.92) | | 7.67 (3.67) | | |  |  | |  |  | |
|  | Perseverative | Sham tDCS | | 14.9 (4.6) | | 11.8 (4.8) | | 13.3 (6.44) | | | 4.10 | .05 (.10) | | 1.52 | .22 (.26) | |
|  |  | Anodal tDCS | | 14.1 (4.8) | | 14.0 (4.56) | | 13.5 (4.6) | | |  |  | |  |  | |
| ***List sort WM*** | Total Score | Sham tDCS | | 31.1 (11.6) | | 31.7 (14.9) | | 34.5 (14.9) | | | 1.24 | .27 (.30) | | **8.12** | **.01 (.02)** | |
|  |  | Anodal tDCS | | 19.9 (13.5) | | 23.7 (14.3) | | 29.2 (18.4) | | |  |  | |  |  | |
| ***Letter Fluency*** | % Correct | Sham tDCS | | 93.4 (7.3) | | 96.9 (3.2) | | 97.4 (4.3) | | | **9.83** | **.003 (.04)** | | **18.65** | **<.001 (.01)** | |
|  |  | Anodal tDCS | | 91.3 (8.5) | | 94.4 (5.0) | | 96.7 (3.8) | | |  |  | |  |  | |
| ***Semantic Fluency*** | % Correct | Sham tDCS | | 94.9 (7.0) | | 96.8 (4.6) | | 97.8 (3.44) | | | 3.27 | .08 (.12) | | 2.00 | .16 (.21) | |
|  |  | Anodal tDCS | | 95.2 (5.0) | | 96.5 (3.3) | | 95.2 (6.4) | | |  |  | |  |  | |
| ***Speed of Processing*** | MRT | Sham tDCS | | 368.6 (34.9) | | 376.0 (39.6) | | 354.8 (34.3) | | | **8.92** | **.004 (.02)** | | 4.58 | .04 (.07) | |
|  |  | Anodal tDCS | | 381.8 (48.1) | | 400.6 (53.0) | | 374.6 (45.0) | | |  |  | |  |  | |
| ***Response Variability*** | CV | Sham tDCS | | .27 (.1) | | .27 (.1) | | .24 (.1) | | | 4.24 | .05 (.12) | | **33.80** | **<.001 (.004)** | |
|  |  | Anodal tDCS | | .31 (.1) | | .29 (.1) | | .28 (.1) | | |  |  | |  |  | |
| ***Prematurity*** | Premature  Responses | Sham tDCS | | 24.24 (46.64) | | 14.28 (25.47) | | 7.39 (2.53) | | | 1.44 | .24 (.29) | | **12.45** | **.001 (.003)** | |
|  |  | Anodal tDCS | | 24.36 (25.37) | | 24.20 (4.82) | | 8.82 (1.75) | | |  |  | |  |  | |
| ***Conners 3-P*** | ADHD Index | Sham tDCS | | 14.4 (3.9) | | 7.7 (4.0) | | 11.1 (5.1) | | | **42.82** | **<.001 (.01)** | | **25.95** | **<.001 (.004)** | |
|  |  | Anodal tDCS | | 16.3 (4.0) | | 12.8 (4.7) | | 11.9 (5.9) | | |  |  | |  |  | |
| ***ARI*** | Parent | Sham tDCS | | .83 (.5) | | .49 (.4) | | .67 (.5) | | | **9.72** | **.003 (.004)** | | 5.14 | .03 (.06) | |
|  |  | Anodal tDCS | | .92 (.6) | | .80 (.6) | | .73 (.5) | | |  |  | |  |  | |
|  | Child | Sham tDCS | | .64 (.5) | | .48 (.52) | | .54 (.5) | | | **12.37** | **.001 (.003)** | | 3.23 | .08 (.12) | |
|  |  | Anodal tDCS | | .82 (.5) | | .64 (.4) | | .67 (.5) | | |  |  | |  |  | |
| ***MEWS*** |  | Sham tDCS | | 16.3 (8.3) | | 14.7 (9.4) | | 16.2 (9.5) | | | 2.38 | .13 (.13) | | .71 | .40 (.40) | |
|  |  | Anodal tDCS | | 18.7 (7.5) | | 17.7 (9.0) | | 16.9 (9.0) | | |  |  | |  |  | |
| ***WREMB-R*** |  | Sham tDCS | | 20.8 (5.3) | | 14.7 (7.3) | | n/a | | | **33.79** | **<.001 (.002)** | |  |  | |
|  |  | Anodal tDCS | | 23.6 (5.6) | | 18.8 (7.3) | | n/a | | |  |  | |  |  | |
| ***CIS*** |  | Sham tDCS | | 21.8 (7.8) | | 24.7 (9.3) | | n/a | | | **14.13** | **<.001 (.002)** | |  |  | |
|  |  | Anodal tDCS | | 16.4 (7.2) | | 20.3 (10.7) | | n/a | | |  |  | |  |  | |
| *repeated measures ANCOVA testing group differences at posttreatment and follow-up (where applicable), adjusting for baseline, age at entry and medication status (naïve, off-medication, on-medication)  ***†***Benjamini-Hochberg adjustment was applied to p-values for time, group, and time by group interaction effect separately and was applied separately to primary cognitive, secondary cognitive, and secondary clinical outcome measures separately. | | | | | | | | | | | | | | | | |

| Table 10. Summary of adjusted averages for ACTIVATE^TM^ (*Magic Lens, Peter’s Printer Panic, Treasure Trunk* only) and Stop Task for each week. Benjamini-Hochberg adjusted p-values in parentheses. | | | | | | | | |
| --- | --- | --- | --- | --- | --- | --- | --- | --- |
|  | ***Adjusted Mean (SD)*** | |  |  | **ANCOVA^a^** | |  |  |
|  | **Sham tDCS** | **Anodal tDCS** | **Time** | | **Group** | | **Time by Group** | |
|  |  |  | ***F (2,88)*** | ***p†*** | ***F (1,44)*** | ***p†*** | ***F (2,88)*** | ***p†*** |
| ***ACTIVATE^TM^*** |  |  |  |  |  |  |  |  |
| Week 1 | 6.67 (.18) | 6.19 (.19) | 2.33 | .10 (.20) | 1.79 | .19 (.19) | .77 | .47 (.47) |
| Week 2 | 14.85 (.42) | 13.89 (.44) |  |  |  |  |  |  |
| Week 3 | 22.91 (.73) | 21.74 (.76) |  |  |  |  |  |  |
|  |  |  | ***F (2,90)*** | ***p*** | ***F (1,44)*** | ***p*** | ***F (2,90)*** | ***P*** |
| ***Stop task*** |  |  | < .001 | 1.00 (1.00) | 2.91 | .10 (.20) | 3.19 | .046 (.09) |
| Week 1 | 49.70 (1.40) | 47.59 (1.46) |  |  |  |  |  |  |
| Week 2 | 50.29 (1.81) | 47.57 (1.89) |  |  |  |  |  |  |
| Week 3 | 54.06 (1.79) | 47.53 (1.86) |  |  |  |  |  |  |
| ^a^Repeated measures ANCOVA testing group differences across the three weeks, adjusting for baseline, age at entry and medication status (Naïve, Off-medication, On-medication), and total game play  †Benjamini-Hochberg adjustment was applied to p-values for time, group, and time by group interaction effects separately and was applied separately to primary cognitive, secondary cognitive, and secondary clinical outcome measures separately. | | | | | | | | |

| Table 11. Summary of Pearson correlations of change scores on CT performance (week 3 minus week 1) correlated with change scores (posttreatment or follow-up minus baseline) for outcomes which showed a significant time effect. Benjamini-Hochberg adjusted p-values in parentheses. | | | | | | | |
| --- | --- | --- | --- | --- | --- | --- | --- |
|  |  |  | | **CT Performance** | | **Stop (%PI)** | |
| **Timepoint Difference** | | |  | ***r*** | ***p†*** | ***r*** | ***p†*** |
| ***Cognitive Measures*** | | |  |  |  |  |  |
| Posttreatment minus Baseline | *Primary Outcome* | |  |  |  |  |  |
|  | GNG Task | | % PI | -0.19 | 0.18 | 0.06 | 0.66 |
|  | *Secondary Outcomes* | |  |  |  |  |  |
|  | Simon Task | | RT Effect | 0.07 | 0.65 (.65) | -0.10 | 0.47 (.82) |
|  | Timing Task | | % Total Correct | 0.23 | 0.12 (.84) | -0.12 | 0.43 (1) |
|  | Mackworth Task | | % Omissions | -0.11 | 0.43 (.60) | -0.22 | 0.12 (.84) |
|  | WCST | | Perseverate Errors | 0.09 | 0.52 (.61) | 0.12 | 0.40 (1) |
|  | Letter Fluency Task | | % Correct | -0.21 | 0.15 (.53) | 0.03 | 0.82 (.96) |
|  | GNG, Simon, CPT | | MRT | -0.16 | 0.28 (.65) | -0.01 | 0.94 (.94) |
|  | GNG, Simon, CPT | | CV | 0.14 | 0.34 (.60) | 0.09 | 0.52 (.73) |
| Follow-Up minus Baseline | *Primary Outcomes* | |  |  |  |  |  |
|  | GNG Task | | % PI | 0.01 | 0.94 (.94) | -0.13 | 0.36 (1) |
|  | CPT | | % Commissions | 0.12 | 0.42 (.63) | 0.07 | 0.64 (.96) |
|  | CPT | | % Omissions | 0.30 | 0.03 (.09) | -0.07 | 0.64 (.64) |
|  | *Secondary Outcomes* | |  |  |  |  |  |
|  | Simon Task | | RT Effect | 0.02 | 0.89 (.89) | -0.02 | 0.88 (.88) |
|  | Mackworth Task | | Omissions | 0.22 | 0.12 (.42) | -0.16 | 0.26 (.91) |
|  | WM | | Total Score | 0.03 | 0.83 (.97) | -0.08 | 0.57 (.80) |
|  | Letter Fluency | | % Correct | -0.30 | 0.04 (.28) | 0.03 | 0.86 (1) |
|  | GNG, Simon, CPT | | MRTS | 0.04 | 0.79 (1) | -0.20 | 0.17 (1) |
|  | GNG, Simon, CPT | | CV | 0.13 | 0.37 (.86) | 0.16 | 0.43 (.75) |
|  | GNG, Simon, CPT | | Premature Responses | 0.09 | 0.52 (.91) | 0.14 | 0.33 (.77) |
| ***Clinical Measures*** | | |  |  |  |  |  |
| Posttreatment minus Baseline | *Primary Outcome* | |  |  |  |  |  |
|  | ADHD-RS | | Total Score | -0.15 | 0.30 | -0.12 | 0.43 |
|  | *Secondary Outcomes* | |  |  |  |  |  |
|  | Conners 3-P | | ADHD Index | -0.18 | 0.21 (.53) | -0.14 | 0.35 (.58) |
|  | ARI | | Parent Score | -0.20 | 0.16 (.80) | -0.20 | 0.16 (.40) |
|  | ARI | | Child Score | -0.08 | 0.58 (.58) | -0.31 | 0.03 (.15) |
|  | CIS | | Total Score | -0.13 | 0.37 (.46) | 0.06 | 0.81 (.81) |
|  | WREMB-R | | Total Score | -0.17 | 0.23 (.38) | -0.09 | 0.56 (.70) |
| Follow-Up minus Baseline | *Primary Outcome* | |  |  |  |  |  |
|  | ADHD-RS | | Total Score | -0.002 | 0.99 | 0.05 | 0.75 |
|  | *Secondary Outcomes* | |  |  |  |  |  |
|  | Conners 3-P | | ADHD Index | -0.09 | 0.55 (.55) | -0.06 | 0.66 (.99) |
|  | ARI | | Parent-Score | -0.18 | 0.20 (.60) | -0.09 | 0.55 (1) |
|  | MEWS | | Total Score | -0.13 | 0.39 (.59) | -0.06 | 0.69 (.69) |
| ***†***Benjamini-Hochberg adjustment was applied to p-values for time, group, and time by group interaction effect separately and was applied separately to primary cognitive, secondary cognitive, and secondary clinical outcome measures separately. | | | | | | | |

| Table 12. Parent and participant ratings of the effectiveness of tDCS and cognitive training split into feedback subscales. | | | | |
| --- | --- | --- | --- | --- |
|  | ***Adjusted Mean (SD)*** | | **ANCOVA^a^** | |
|  | **Sham tDCS** | **Anodal tDCS** | ***F (1, 45)*** | ***p*** |
| **Parent** |  |  |  |  |
| *Liking* | 3.07 (.88) | 2.89 (.88) | .48 | .49 |
| *Effectiveness* | 3.26 (.81) | 3.03 (.81) | .96 | .33 |
| *Recommend* | 3.6 (.87) | 3.30 (.87) | 1.42 | .24 |
| **Child** |  |  |  |  |
| *Learning* | 3.55 (.73) | 3.40 (.75) | .49 | .49 |
| *Transfer* | 3.12 (.62) | 2.87 (.65) | 1.67 | .20 |
| *Recommend* | 3.92 (.79) | 3.61 (.81) | 1.18 | .19 |
| *Liking* | 3.73 (1.11) | 3.62 (1.11) | .12 | .73 |
| ^a^univariate ANCOVA testing group differences for each feedback rating subscale separately, covarying for age in months and medication status; | | | | |

| Table 13. Adjusted weekly averages of participant-ratings for mood and wakefulness before and during tDCS & CT sessions over the treatment trial period. | | | | | | | | |
| --- | --- | --- | --- | --- | --- | --- | --- | --- |
|  | ***Adjusted Mean (SD)*** | | **ANCOVA^a^** | | | | | |
|  | **Sham tDCS** | **Anodal tDCS** | **Time** | | **Group** | | **Time by Group** | |
|  |  |  | ***F*** | ***p*** | ***F*** | ***p*** | ***F*** | ***p*** |
| **Mood before** |  |  |  |  |  |  |  |  |
| *Week 1* | 3.58 (.68) | 3.58 (.68) | 0.97 | 0.38 | 0.75 | 0.39 | 1.71 | 0.57 |
| *Week 2* | 3.52 (.90) | 3.81 (.90) |  |  |  |  |  |  |
| *Week 3* | 3.58 (.79) | 3.83 (.79) |  |  |  |  |  |  |
| **Mood during** |  |  |  |  |  |  |  |  |
| *Week 1* | 3.71 (.72) | 3.86 (.73) | 0.18 | 0.85 | 0.87 | 0.36 | 0.89 | 0.35 |
| *Week 2* | 3.67 (.97) | 3.77 (.97) |  |  |  |  |  |  |
| *Week 3* | 3.64 (.71) | 3.97 (.71) |  |  |  |  |  |  |
| ^a^repeated measures ANCOVA testing group differences across the three weeks of the treatment trial period, covarying for age in months and medication status | | | | | | | | |

| Table 14. Summary of the differences of tolerability of skin sensations generated by anodal and sham tDCS and their effect on cognitive training performance. | | | | |
| --- | --- | --- | --- | --- |
|  | **Mean (SD)** | | **ANOVA** | |
|  | **Sham tDCS** | **Anodal tDCS** | **F (1,45)** | ***p*** |
| **Have you experienced any sensation during the stimulation (1 = none, 2 = mild, 3 = moderate; 4 = considerable; 5 = strong)?** | | | | |
| *Itching* | .97 (.14) | .95 (.15) | .004 | .93 |
| *Pain* |  |  | .98 | .33 |
| *Burning* | .21 (.13) | .62 (.14) | 4.20 | **.05** |
| *Warmth/heat* | .43 (.13) | .32 (.12) | .34 | .57 |
| *Pinching* | .28 (.08) | .20 (.09) | .37 | .55 |
| *Metallic* | .06 (.07) | .16 (.07) | .86 | .36 |
| *Fatigue* | .27 (.13) | .43 (.14) | .70 | .41 |
|  | ***N*** | |  |  |
| **When did the discomfort begin?** |  |  | **χ^2^ (3)** | ***p*** |
| *n/a* | 3 | 1 | 3.37 | .34 |
| *Beginning* | 16 | 20 |  |  |
| *Middle* | 6 | 3 |  |  |
| *End* | 1 | 0 |  |  |
| **How long did it last? It stopped…** |  |  | **χ^2^ (3)** | ***p*** |
| n/a | 3 | 1 | .67 | .88 |
| …quickly | 16 | 20 |  |  |
| …in the middle | 6 | 3 |  |  |
| …at the end | 1 | 0 |  |  |
| **Did you find that the sensations you felt affected how you played the games?** |  |  | **χ^2^ (3)** | ***p*** |
| Yes | 3 | 1 | 1.27 | .26 |
| No | 21 | 25 |  |  |
| **How much did it affect your performance (1 = not at all, 2 = slightly, 3 = considerably; 4 = much; 5 = very much)?** | | | **χ^2^ (3)** | ***p*** |
|  | 1.13 (.13) | 1.41 (.13) | 2.27 | .14 |
